# Calorie reformulation: A systematic review and meta-analysis examining the effect of manipulating food energy density on daily energy intake and body weight

**DOI:** 10.1101/2021.11.04.21265933

**Authors:** Eric Robinson, Mercedes Khuttan, India McFarland-Lesser, Zina Patel, Andrew Jones

**Affiliations:** Department of Psychology, Eleanor Rathbone Building, University of Liverpool Liverpool, L69 7ZA, UK

**Keywords:** energy density, energy intake, obesity, food reformulation, low fat

## Abstract

**Background:** Dietary energy density is thought to be a contributor to obesity, but the extent to which different magnitudes and types of reductions to food energy density decreases daily energy intake is unclear.

**Objective:** To systematically review and meta-analyse experimental studies that have examined the effect that manipulating energy density of food has on total daily energy intake.

**Design:** A systematic review and multi-level meta-analysis of studies on human participants that used an experimental design to manipulate the energy density of foods served and measured energy intake for a minimum of one day.

**Results:** Thirty-one eligible studies contributed 90 effects comparing the effect of higher vs. lower energy density of served food on daily energy intake to the primary meta-analysis. Lower energy density of food was associated with a large decrease in daily energy intake (SMD = -1.002 [95% CI: -0.745 to -1.266]). Findings were consistent across studies that did vs. did not manipulate macronutrient content to vary energy density. The relation between decreasing energy density and daily energy intake tended to be strong and linear, whereby compensation for decreases to energy density of foods (i.e. by eating more at other meals) was minimal. Meta-analysis of (n=5) studies indicated that serving lower energy dense food tended to be associated with greater weight loss than serving higher energy dense food, but this difference was not significant (−0.7kg, 95% CIs: -1.34, 0.04).

**Conclusions:** Decreasing the energy density of food can substantially reduce daily energy intake and may therefore be an effective public health approach to reducing population level energy intake.

## Introduction

Energy density is the amount of energy contained in a given weight of food (kcal/gram). Fat (9kcal/g), protein (4kcal/g), carbohydrate (4kcal/g) and water (0kcal/g) content explain variations in food energy density (1). Although some foods tend to be higher in energy density (e.g. confectionary) than others (e.g. fruits and vegetables), even among the same type of food product there can be substantial variation in energy density between product brands (2). Increased availability of low-cost energy dense food products is likely a major contributor to higher obesity prevalence (3-5). Epidemiological data indicate that diets which are more energy dense tend to be associated with higher daily energy intake and weight gain (6-9). These findings have led to suggestions that population level approaches to reduce dietary energy density may be effective in reducing obesity (1, 10, 11). However, epidemiological studies examining energy density and risk of obesity have produced some inconsistent findings (12, 13), which may be due to the methodological challenges of accurately measuring dietary energy density and daily energy intake from self-report measures (6, 7, 13).

A number of laboratory studies have examined the causal impact that food energy density has on short-term energy intake, and it is now well established that reducing the energy density of a meal decreases acute energy consumed at that meal (10, 14). The longer-term effects of manipulating food energy density on energy intake are less well understood (14). Studies which have manipulated the energy density of some, or all food served to participants throughout the day indicate that daily energy intake tends to decrease when the energy density of served food is reduced (15-20). However, as energy density may influence the physiological processing and digestion of food (14), consumers may at least in part ‘compensate’ for reductions to food energy density by increasing consumption of other food. In line with this, there is some evidence that self-reported hunger is higher after consumption of lower energy dense food as opposed to higher energy dense foods (21). When weight of food is held constant lower energy dense foods are also associated with greater later food intake than higher energy dense foods (22). From an applied perspective, it will be important to understand whether the method used to alter energy density of food, such as holding macronutrient composition constant vs. altering macronutrient composition (e.g. reducing % of kcals from fat), has a significant impact on the extent to which consumers compensate for changes in food energy density (19). It is also possible that the impact reducing energy density has on energy intake may be non-linear. In particular, compensatory responses in appetite may be more likely to occur for foods that are lower in energy density (i.e. < 1.75kcal/g), as opposed to more highly energy dense foods (23), as it has been suggested that humans evolutionary past leaves them poorly adapted to the recent emergence of very energy dense foods (24). Yet, these questions remain unanswered in relation to the impact that manipulating food energy density has on daily energy intake. In addition, although there is some evidence that dietary advice designed to decrease dietary energy density may benefit weight loss (1, 25), there is currently a lack of consensus on the direct impact of reformulating the energy density of food products has on body weight.

The primary aim of the present research was to systematically review and meta-analyse studies that have examined the impact that reducing energy density of served food has on daily energy intake. Secondary aims included understanding moderators of the effect that altering energy density has on daily energy intake and effects on body weight.

## Method

PRISMA guidelines were followed (26). This review was registered on PROSPERO (CRD42020223973) and the analysis protocol was pre-registered https://osf.io/dj4yf/ In the pre-registration we intended to review studies on both energy density and portion size in the same report. However, prior to data extraction we updated the protocol to review portion size and energy density studies separately due to the large number of studies identified. Here we focus on energy density studies and studies on portion size are reviewed elsewhere (27).

### Eligibility criteria

#### Participants

Only studies sampling human participants were eligible. Studies were excluded that sampled participants who were currently undergoing any medical treatment which may influence appetite (e.g. bariatric surgery).

#### Intervention

Studies were required to have manipulated the energy density of food products or meals (i.e. energy content divided by weight of food served; kcal/gram) served to participants. Studies were included that manipulated the energy density of a minimum of one food/meal, and studies that manipulated energy density of up to all foods/meals served across the day were eligible. If a study only manipulated the energy density of a beverage it was deemed ineligible, as the main focus was on food energy density. However, if a study manipulated the energy density of multiple foods/meals and also extended the manipulation to accompanying drinks, it was eligible. To be eligible, studies were required to manipulate energy density by serving participants one or more varying energy densities of the same or very similar type of food/meal (e.g. lower vs. higher energy dense tomato-based pasta dish). Eligible manipulations of energy density included altering the % of energy derived from fat, protein and/or carbohydrate (e.g. standard vs. low-fat cheese). Studies that manipulated energy density through altering water content (e.g. adding water to a porridge) or substitution of lower energy dense foods (e.g. vegetables) were eligible.

#### Comparator

In studies with two energy density conditions the ‘comparator’ condition was the condition with the highest energy density and the condition with the lowest energy density was the ‘intervention’ condition (as public health interventions tend to aim to decrease energy intake). It was common for studies to have multiple energy density conditions (e.g. higher vs. medium vs. lower) and all contrasts were included for use as individual contrasts (e.g. higher vs. medium, higher vs. lower, medium vs. lower).

#### Outcomes

To be eligible, studies had to have measured energy intake for a minimum of one day (i.e. at least 3 main meals). Measurements of energy intake that were based on an objective researcher measurement (e.g. weighing of food pre/post eating in the laboratory), participant self-reports (e.g. dietary recall) or a combination were eligible. Energy intake could be assessed under controlled laboratory settings or in real-world settings to be eligible. Measures of energy intake that were not determined by sampled participants (e.g. an infant being bottle or spoon fed) were not eligible.

#### Study Design

Studies using a within-subjects/repeated measures design (i.e. participants receive all energy density conditions) or a between-subjects design studies (i.e. participants receive only one energy density condition) were eligible. Some studies required participants to consume a meal/food in full (e.g. consumption of a set amount of energy density manipulated food) and these designs were eligible. Studies that ‘crossed’ energy density manipulations with another experimental factor (e.g. manipulation of both energy density of food and portion sizes in the same study) were eligible. For studies that did not manipulate energy density of all meals/foods, studies were required to measure and report energy intake at that meal(s) that energy density was manipulated for in order to be eligible (to permit quantification of the effect of the energy density manipulation independent of non-manipulated foods/meals).

### Search process and article identification

The electronic databases PsycINFO, PubMed and SCOPUS (from date of inception) were searched during September-October 2020. For combinations of search terms used please refer to online supplementary material. The reference lists of all eligible papers were searched and also contacted authors of included studies to inquire about any further eligible studies. Potential grey literature was addressed (to minimize publication bias) by searching the OSF preprint archive (includes 30 other preprint archives, including Nutrixiv). Two authors independently screened and judged eligibility of all articles identified through electronic searches. One author completed the snowballing and grey literature searching approaches to identify any additional potentially eligible articles, eligibility was confirmed by a second author. Any discrepancies during eligibility assessments were discussed with a third author. Searches were re-run in October 2021 to identify any new eligible studies published between 2020-2021. None were identified.

### Data extraction

The following study information was extracted by two authors independently (any discrepancies were resolved through discussion or by a third author); sampled participants (e.g. country, participant group sampled, summary information concerning sample demographics), energy density manipulation (e.g. number of foods/meals manipulated, energy density in each condition (kcal/g), nutritional composition (energy from protein, fat and carbohydrates) of energy density conditions, total number of kcals served in energy density conditions (if reported), design of study (e.g. within or between-subjects), energy intake measure (self-report vs. researcher measured, vs. mixed), whether any foods/meals had to be eaten in full (compulsory eating vs. ad libitum), number of days energy intake was measured for, energy intake information (e.g. energy intake under different conditions of energy density), whether body weight was measured before and after the different energy density conditions, and study factors related to risk of bias (see below). Authors were contacted and asked to provide details if statistical information required for analyses examining energy intake was missing (e.g. standard error not reported for energy intake under different energy density conditions).

### Risk of bias ratings

A risk of bias checklist was created from existing study quality assessment tools and best practice recommendations for studying energy intake under experimental conditions (28-32), as existing bias tools (e.g. Cochrane) were not relevant to studies examining the effect of experimental manipulations of energy density on energy intake. The checklist included nine items for extraction and ‘yes’ was indicative of higher risk of bias; Was measured energy intake dependent on participant self-reporting? Did the study fail to use key participant exclusion criteria (e.g. use of medication affecting appetite)? Was any key methodological detail missing (e.g. limited information on procedures)? Was a non-random method of allocation to the different energy density condition used allocation (or was allocation method not described)? Were participants required to consume any study foods/meals in their entirety? Were demand characteristics not addressed in the study (e.g. no attempt to blind participants to study aims or measure whether differences between energy density foods were detectable)? Did the study have a small sample size (N<12 for within-subject designs)? Was the study pre-registered? Was there an absence of information on conflicts of interest or a reported relevant conflict of interest?

### Analyses

The pre-registered analysis protocol and data are available online at: https://osf.io/dj4yf/. Deviations from planned analyses are reported in the online supplemental material.

#### Primary analyses

##### Effect of energy density condition on daily energy intake

We first examined the effect of energy density condition on daily energy intake. Because a number of studies contributed multiple energy density comparisons (e.g. lower, medium, higher), we used multi-level meta-analysis (33). Studies did not report on the correlation between daily energy intake under the different energy density conditions and we therefore imputed the size of this correlation based on similar studies (27) of daily energy intake (r = 0.8)) and we conducted sensitivity analyses varying magnitude (0.6, 0.4) to examine consistency of results. Outliers were identified as effect sizes which the upper bound of their 95% confidence interval was lower than the lower bound of the meta-analysed pooled effect confidence interval of all effects or for which the lower bound of their 95% confidence interval was higher than the meta-analysed pooled effect confidence interval of all effects. Influential cases were identified as any effects with DFBETA values > 1 (indicative of a >1 change in the standard deviation of the estimated co-efficient after removal) (34) and we also conducted leave one out analyses. Egger’s test for publication bias (35) and the trim and fill procedure for funnel plot asymmetry (36) were used. More detailed information is available in the online supplementary materials. If any outliers were identified, we examined the effect of removing them from the main primary meta-analysis. We also excluded them from our subsequent primary sub-group and meta-regression analyses on daily energy intake to minimize results being driven by large effects, but also examined if results were consistent when included. Dependent on outcome of interest, we either report meta-analyses as standardised mean difference (SMD), whereby SMDs of 0.2, 0.5 and 0.8 are small, moderate, and large statistical sized effects retrospectively (37) or report the mean weighted difference in energy intake (kcals) between energy density conditions to aid interpretation.

##### Sub-group and meta-regression analyses

To examine whether participant or study features moderated the effect of energy density condition on daily energy intake, we conducted a series of sub-group analyses and meta-regressions. For sub-group analyses, a-priori a minimum of n=5 effects per sub-group were required. We examined the effect of age group (children vs. adult samples), sex (female vs. male vs. mixed), number of meals/foods energy density was manipulated for (all meals served vs. not), whether energy density manipulation altered macronutrient composition (% of kcals from protein vs. fat vs. carbohydrates altered between conditions vs. kept constant) and number of days energy intake was assessed for in the study (meta-regression). We also assessed risk of bias indicators for which there was sufficient variability between studies in a series of sub-groups analyses; use of random allocation (yes vs. no/not reported), energy intake measure (objective vs. reliant on self-report), whether demand characteristics were assessed (addressed vs. not addressed), conflicts of interest (statement included and no conflict vs. conflict reported or unclear). Because we found strong evidence that whether a study manipulated all foods/meals (vs did not) had a large impact on daily energy intake, we decided (unplanned) it was more appropriate to examine the relationship between absolute difference in energy density between energy density conditions (meta-regression, expressed as kcal/g) and daily energy intake for the two study types separately (see *Analyses by study* type below).

##### Linearity of relationship between manipulating energy density and energy intake

There was sufficient variability across studies to examine whether changes to energy density occurring at lower energy densities (e.g. reducing energy density of a relatively low energy dense food) produced smaller sized effects on daily energy intake as changes to energy density occurring at higher levels of energy density (e.g. reducing energy density of an energy dense food). We assessed this using both meta-regression (expressed as kcal/g of largest energy density condition) and in line with (23), a sub-group analysis that compared effects in which both energy density conditions < 1.75kcal/g vs. ≥1 energy density conditions exceeded 1.75kcal/g.

##### Analyses by study type

As the impact of manipulating energy density on daily energy intake differed substantially based on whether all meals/foods were manipulated vs. not, we conducted separate analyses for these two study types. For studies manipulating energy density of all food/meals, we examined whether absolute difference in energy density between energy density conditions was associated with daily energy intake (meta-regression), as well as repeating primary analyses examining number of days energy intake was assessed for and linearity of relationship between manipulating energy density and energy intake for sensitivity purposes. Next, we conducted analyses among studies that did not manipulate all foods/meals and also reported on energy intake during both manipulated meals and non-manipulated meals. We repeated the same analyses as above, as well as examining whether compulsory eating (participants required to consume one or meals eaten in full vs. ad libitum consumption) moderated results, as it was only common among this study type. We also examined the relationship between the total difference in kcals served between energy density conditions and daily energy intake using meta-regression. To further aid interpretation of ‘compensation’ effects after consuming lower vs. higher energy density foods/meals, we conducted separate meta-analyses on energy intake from manipulated foods and non-manipulated foods separately. In some instances, the manipulated meal/food was ‘fixed’ (i.e. it was compulsory for participants to eat the meal in full) and this equates to a standard error of 0, entered as 0.1 in meta-analysis. In sensitivity analyses we imputed these values as the average SE (expressed as a proportion of mean energy intake) taken from energy density manipulated meals that were not ‘fixed’, in order to ensure results were consistent.

##### Body weight

A small number of studies (n=5) reported data on change in body weight before vs. after lower and higher energy density conditions. Standard deviations were not reported and therefore we imputed this (based on average SD as a % of M weight change) from (27) and used sensitivity analyses to examine consistency when size of SD was larger and smaller. We used generic variance inverse meta-analysis to pool change in body weight (kg). In instances where a study had multiple energy density conditions, to maximise statistical power, a-priori we only included the energy density condition contrast with the largest difference in energy density served.

## Results

### Summary of included studies

A total of 31 eligible studies were included in the review and meta-analysis (see Table 1). Figure 1 outlines the study selection process. Twenty-seven studies sampled adults and the remaining four studies sampled children. Most included studies were conducted in the US (n=19) and the remainder where in Europe (n=9) or Singapore (n=3). All studies used within-subjects designs to examine the effect of manipulating energy density. Energy intake was assessed between 1 and 14 days in studies and the most common study length was 1 day (n=18). Sample sizes of studies ranged from N=6 to N=95. Twenty-three studies manipulated either a single meal or a limited number of meals/food items (as opposed to all meals/foods) and the remaining (n=8) studies manipulated energy density of all food served to participants. It was most common for studies to compare the effect of two energy density conditions on daily energy intake (n=22), eight studies compared three energy density conditions and a single study had five energy density conditions. For n=9 studies, the macronutrient content (i.e. %kcals from protein, fat, carbohydrates) of food served in the different energy density conditions was held constant. For n=14 studies, energy density was manipulated by altering macronutrient content (e.g. reducing %kcals from fat) and in one study macronutrient content was held constant across two of the density conditions and differed between two energy density conditions. In n=7 studies, macronutrient information was not reported or unclear. The lowest energy density condition in a study was 0.11–0.13kcal/g (the higher energy density condition from this study was 0.49-0.5kcal/g). The highest energy density condition in a study was 5.47kcal/g (the low energy density condition in the study was 2.53kcal/g), this study also had the largest absolute difference (2.94kcal/g). See Table 1 for individual study information.

**Table 1.**
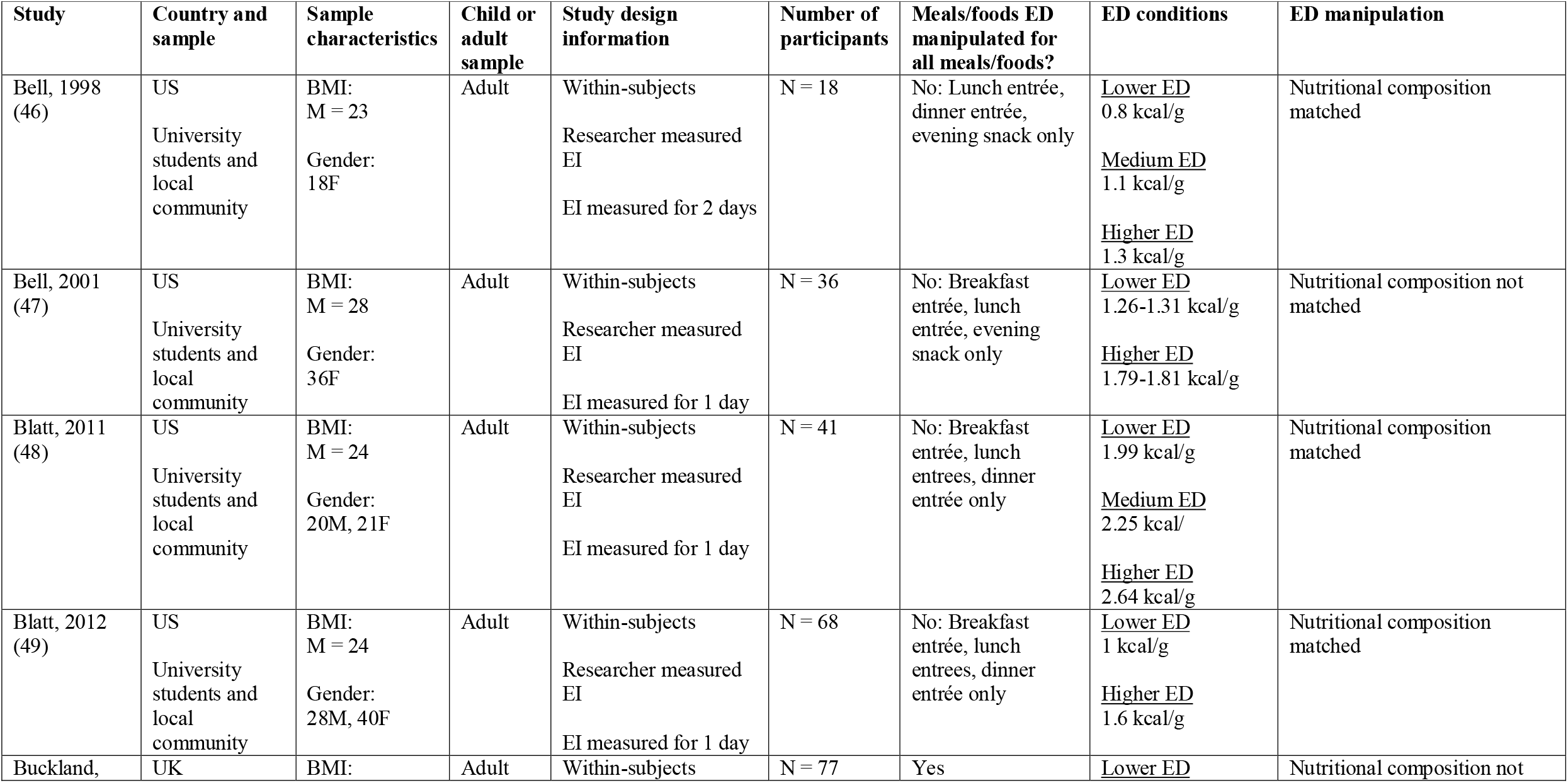
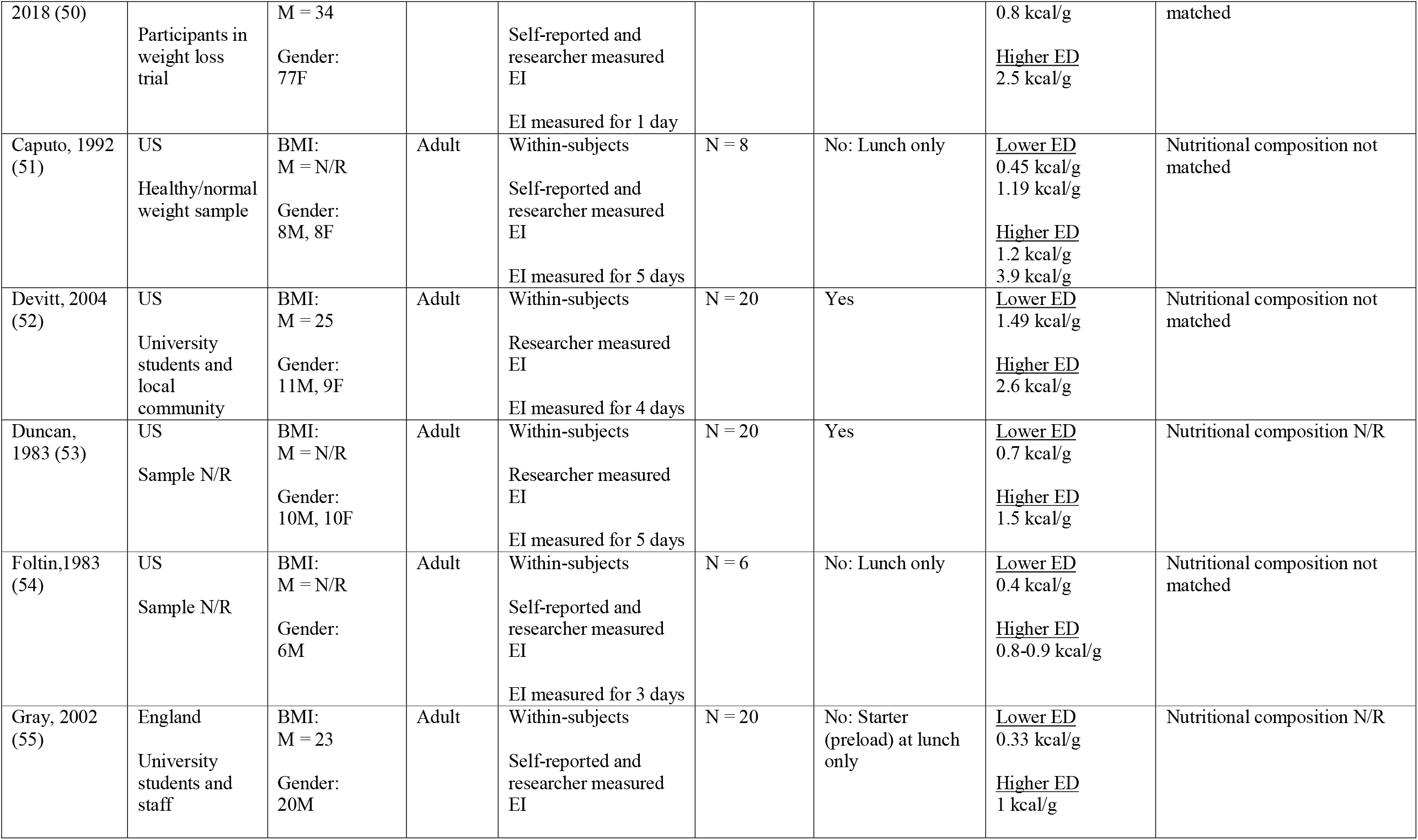
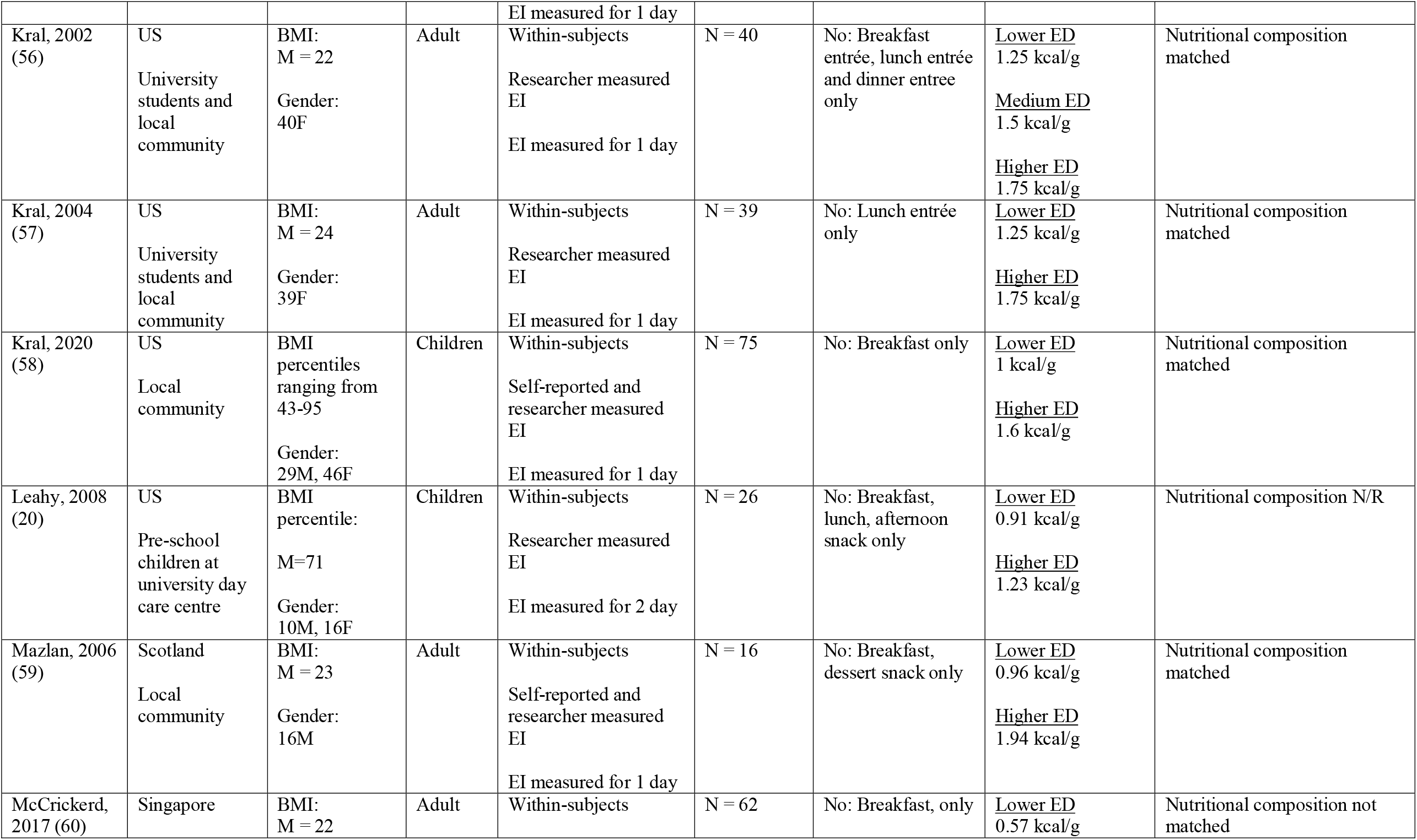
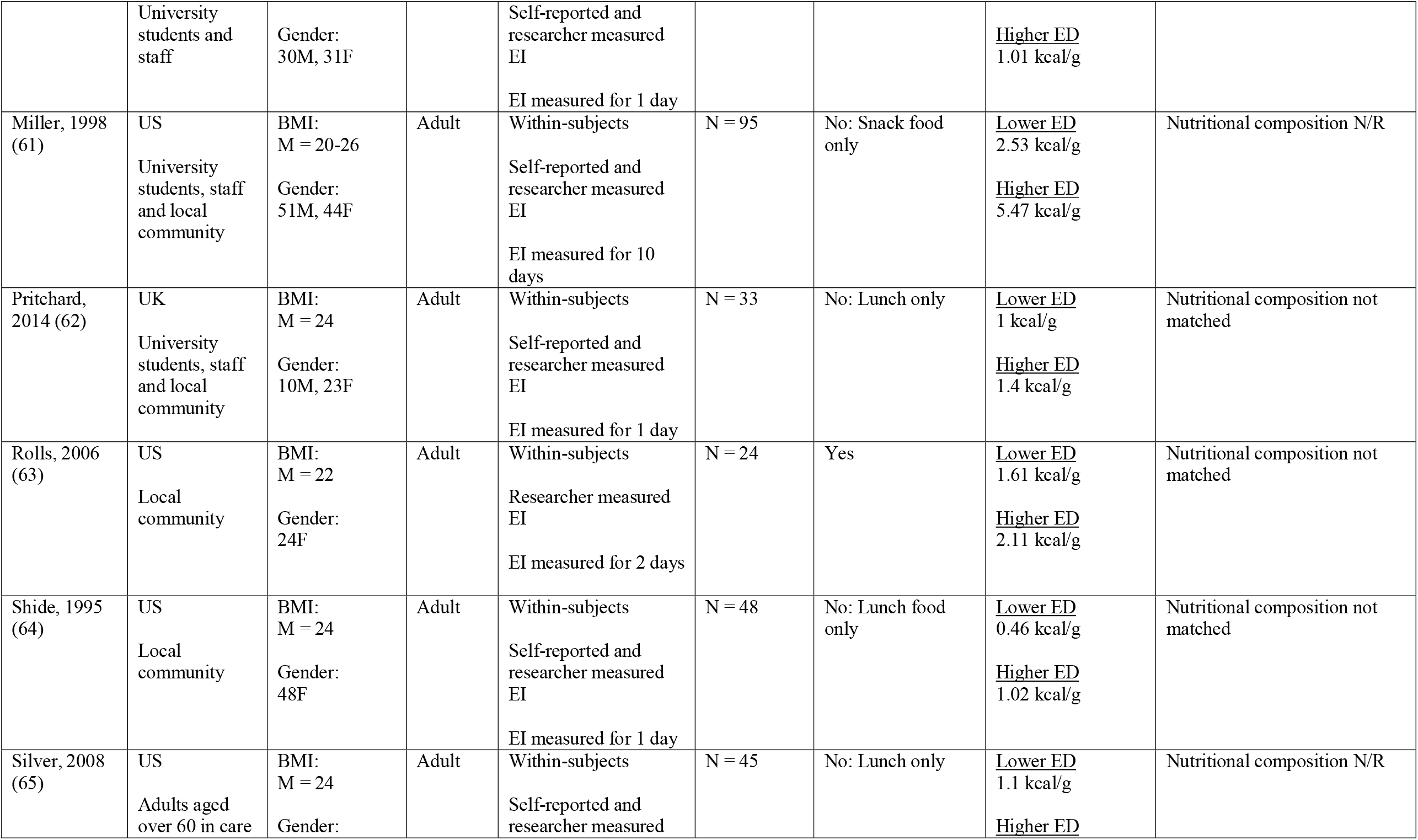
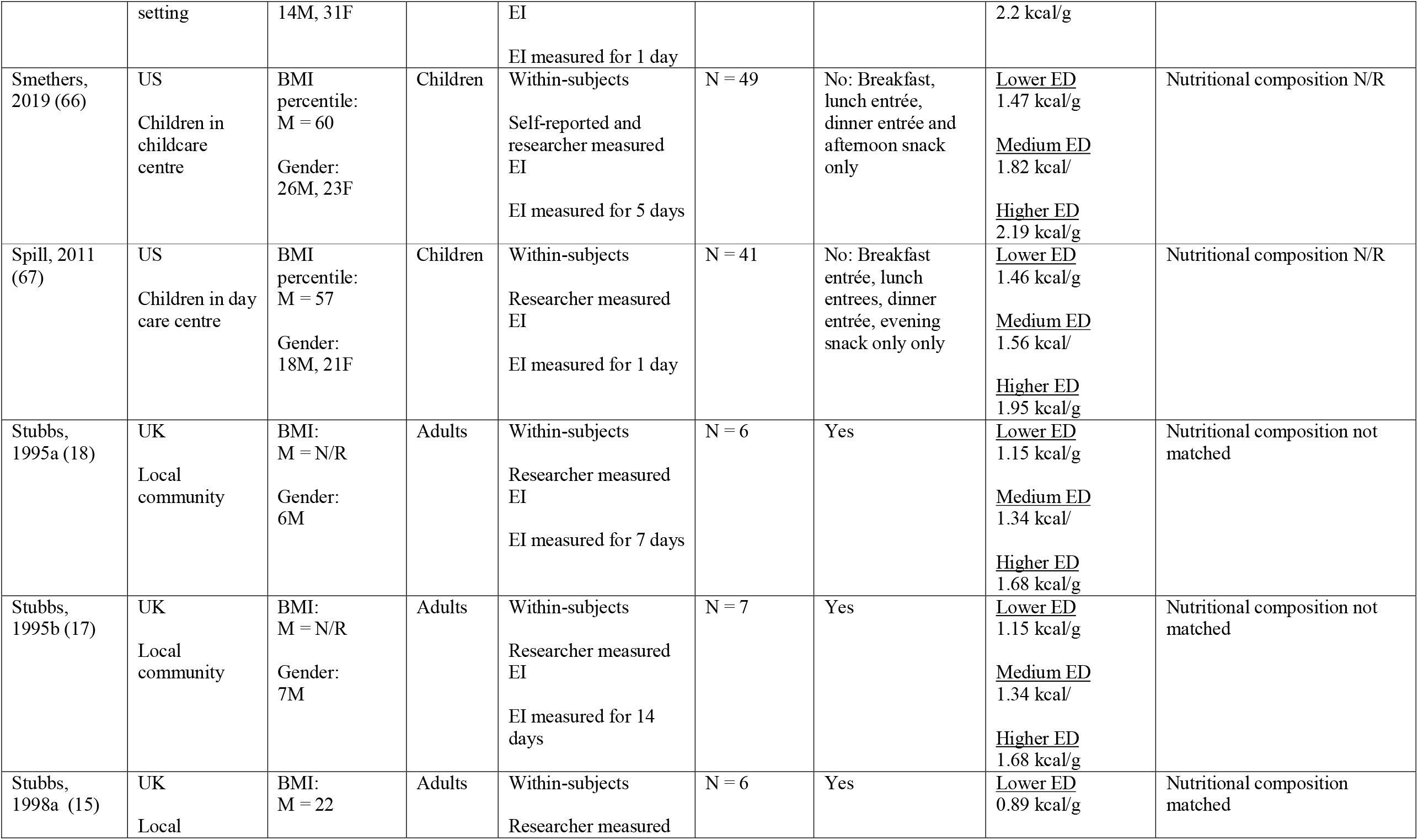
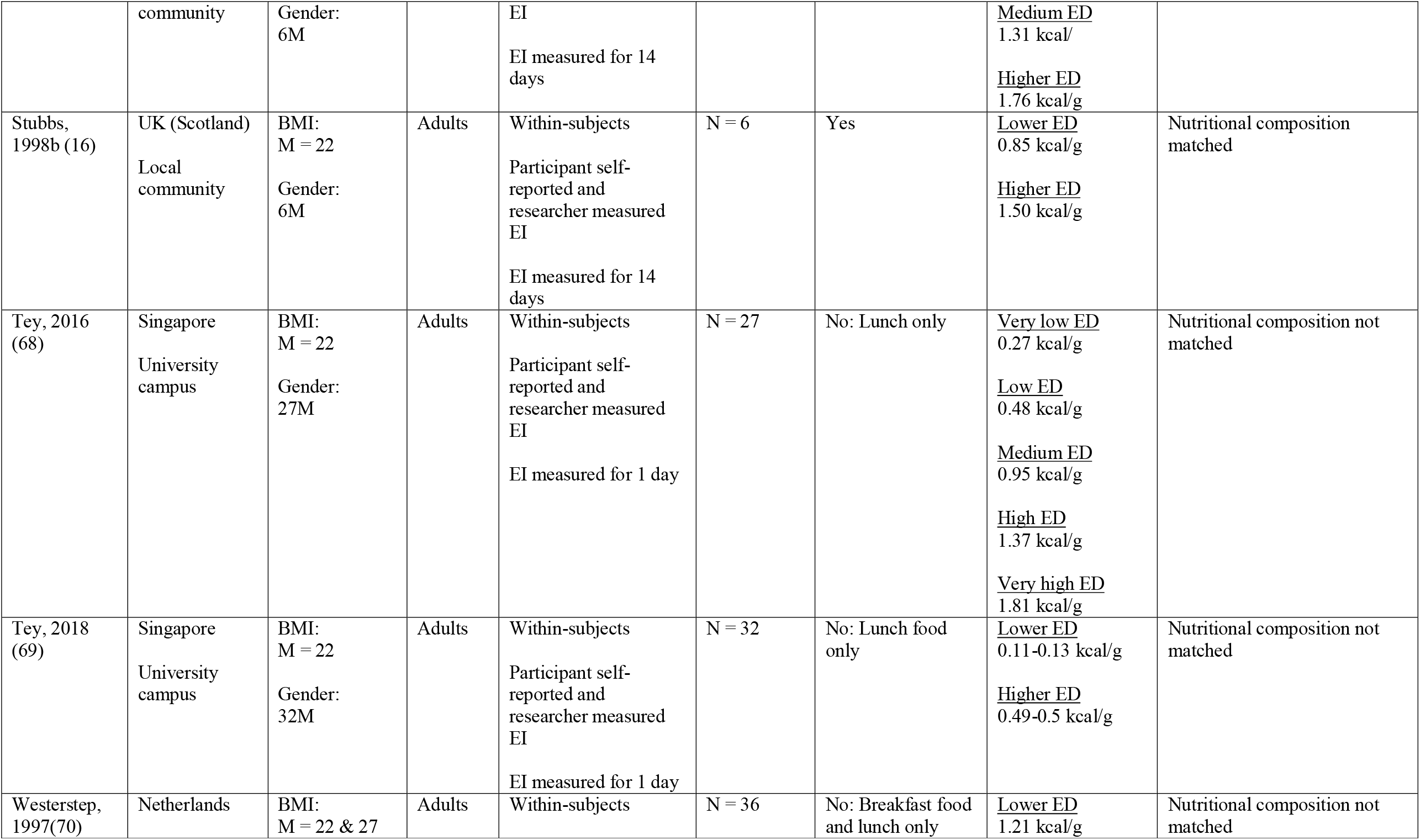
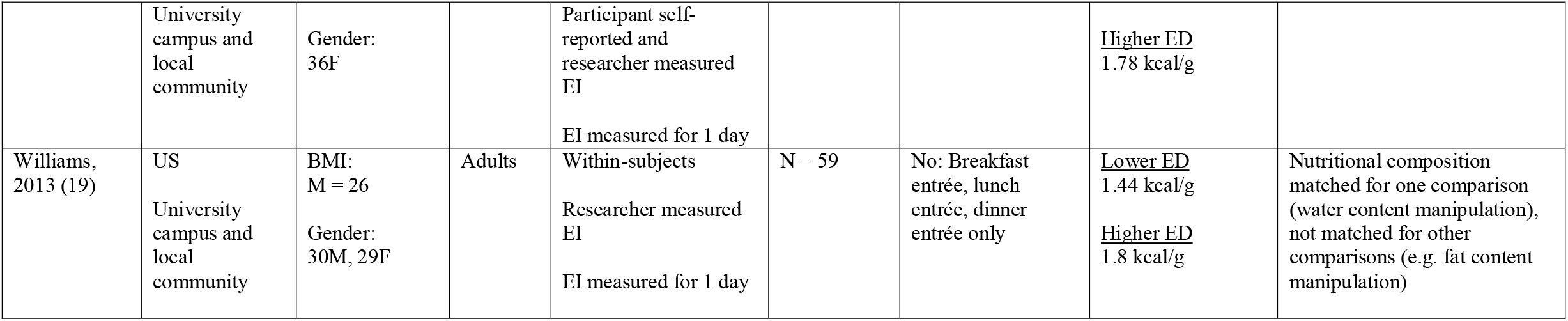
Summary information on included studies

| Study | Country and sample | Sample characteristics | Child or adult sample | Study design information | Number of participants | Meals/foods ED manipulated for all meals/foods? | ED conditions | ED manipulation |
| --- | --- | --- | --- | --- | --- | --- | --- | --- |
| Bell, 1998 (46) | US<br>University students and local community | BMI:<br>M = 23<br>Gender:<br>18F | Adult | Within-subjects<br>Researcher measured EI<br>EI measured for 2 days | N = 18 | No: Lunch entrée, dinner entrée, evening snack only | <u>Lower ED</u><br>0.8 kcal/g<br><u>Medium ED</u><br>1.1 kcal/g<br><u>Higher ED</u><br>1.3 kcal/g | Nutritional composition matched |
| Bell, 2001 (47) | US<br>University students and local community | BMI:<br>M = 28<br>Gender:<br>36F | Adult | Within-subjects<br>Researcher measured EI<br>EI measured for 1 day | N = 36 | No: Breakfast entrée, lunch entrée, evening snack only | <u>Lower ED</u><br>1.26-1.31 kcal/g<br><u>Higher ED</u><br>1.79-1.81 kcal/g | Nutritional composition not matched |
| Blatt, 2011 (48) | US<br>University students and local community | BMI:<br>M = 24<br>Gender:<br>20M, 21F | Adult | Within-subjects<br>Researcher measured EI<br>EI measured for 1 day | N = 41 | No: Breakfast entrée, lunch entrees, dinner entrée only | <u>Lower ED</u><br>1.99 kcal/g<br><u>Medium ED</u><br>2.25 kcal/<br><u>Higher ED</u><br>2.64 kcal/g | Nutritional composition matched |
| Blatt, 2012 (49) | US<br>University students and local community | BMI:<br>M = 24<br>Gender:<br>28M, 40F | Adult | Within-subjects<br>Researcher measured EI<br>EI measured for 1 day | N = 68 | No: Breakfast entrée, lunch entrees, dinner entrée only | <u>Lower ED</u><br>1 kcal/g<br><u>Higher ED</u><br>1.6 kcal/g | Nutritional composition matched |
| Buckland, | UK | BMI: | Adult | Within-subjects | N = 77 | Yes | <u>Lower ED</u> | Nutritional composition not |
| 2018 (50) | Participants in weight loss trial | M = 34<br>Gender: 77F |  | Self-reported and researcher measured EI<br>EI measured for 1 day |  |  | 0.8 kcal/g<br><u>Higher ED</u><br>2.5 kcal/g | matched |
| Caputo, 1992 (51) | US<br>Healthy/normal weight sample | BMI: M = N/R<br>Gender: 8M, 8F | Adult | Within-subjects<br>Self-reported and researcher measured EI<br>EI measured for 5 days | N = 8 | No: Lunch only | <u>Lower ED</u><br>0.45 kcal/g<br>1.19 kcal/g<br><u>Higher ED</u><br>1.2 kcal/g<br>3.9 kcal/g | Nutritional composition not matched |
| Devitt, 2004 (52) | US<br>University students and local community | BMI: M = 25<br>Gender: 11M, 9F | Adult | Within-subjects<br>Researcher measured EI<br>EI measured for 4 days | N = 20 | Yes | <u>Lower ED</u><br>1.49 kcal/g<br><u>Higher ED</u><br>2.6 kcal/g | Nutritional composition not matched |
| Duncan, 1983 (53) | US<br>Sample N/R | BMI: M = N/R<br>Gender: 10M, 10F | Adult | Within-subjects<br>Researcher measured EI<br>EI measured for 5 days | N = 20 | Yes | <u>Lower ED</u><br>0.7 kcal/g<br><u>Higher ED</u><br>1.5 kcal/g | Nutritional composition N/R |
| Foltin, 1983 (54) | US<br>Sample N/R | BMI: M = N/R<br>Gender: 6M | Adult | Within-subjects<br>Self-reported and researcher measured EI<br>EI measured for 3 days | N = 6 | No: Lunch only | <u>Lower ED</u><br>0.4 kcal/g<br><u>Higher ED</u><br>0.8-0.9 kcal/g | Nutritional composition not matched |
| Gray, 2002 (55) | England<br>University students and staff | BMI: M = 23<br>Gender: 20M | Adult | Within-subjects<br>Self-reported and researcher measured EI | N = 20 | No: Starter (preload) at lunch only | <u>Lower ED</u><br>0.33 kcal/g<br><u>Higher ED</u><br>1 kcal/g | Nutritional composition N/R |
|  |  |  |  | EI measured for 1 day |  |  |  |  |
| Kral, 2002 (56) | US<br>University students and local community | BMI:<br>M = 22<br>Gender:<br>40F | Adult | Within-subjects<br>Researcher measured EI<br>EI measured for 1 day | N = 40 | No: Breakfast entrée, lunch entrée and dinner entree only | <u>Lower ED</u><br>1.25 kcal/g<br><u>Medium ED</u><br>1.5 kcal/g<br><u>Higher ED</u><br>1.75 kcal/g | Nutritional composition matched |
| Kral, 2004 (57) | US<br>University students and local community | BMI:<br>M = 24<br>Gender:<br>39F | Adult | Within-subjects<br>Researcher measured EI<br>EI measured for 1 day | N = 39 | No: Lunch entrée only | <u>Lower ED</u><br>1.25 kcal/g<br><u>Higher ED</u><br>1.75 kcal/g | Nutritional composition matched |
| Kral, 2020 (58) | US<br>Local community | BMI percentiles ranging from 43-95<br>Gender:<br>29M, 46F | Children | Within-subjects<br>Self-reported and researcher measured EI<br>EI measured for 1 day | N = 75 | No: Breakfast only | <u>Lower ED</u><br>1 kcal/g<br><u>Higher ED</u><br>1.6 kcal/g | Nutritional composition matched |
| Leahy, 2008 (20) | US<br>Pre-school children at university day care centre | BMI percentile:<br>M=71<br>Gender:<br>10M, 16F | Children | Within-subjects<br>Researcher measured EI<br>EI measured for 2 day | N = 26 | No: Breakfast, lunch, afternoon snack only | <u>Lower ED</u><br>0.91 kcal/g<br><u>Higher ED</u><br>1.23 kcal/g | Nutritional composition N/R |
| Mazlan, 2006 (59) | Scotland<br>Local community | BMI:<br>M = 23<br>Gender:<br>16M | Adult | Within-subjects<br>Self-reported and researcher measured EI<br>EI measured for 1 day | N = 16 | No: Breakfast, dessert snack only | <u>Lower ED</u><br>0.96 kcal/g<br><u>Higher ED</u><br>1.94 kcal/g | Nutritional composition matched |
| McCrickerd, 2017 (60) | Singapore | BMI:<br>M = 22 | Adult | Within-subjects | N = 62 | No: Breakfast, only | <u>Lower ED</u><br>0.57 kcal/g | Nutritional composition not matched |
|  | University students and staff | Gender: 30M, 31F |  | Self-reported and researcher measured EI<br>EI measured for 1 day |  |  | <u>Higher ED</u><br>1.01 kcal/g |  |
| Miller, 1998 (61) | US<br>University students, staff and local community | BMI: M = 20-26<br>Gender: 51M, 44F | Adult | Within-subjects<br>Self-reported and researcher measured EI<br>EI measured for 10 days | N = 95 | No: Snack food only | <u>Lower ED</u><br>2.53 kcal/g<br><u>Higher ED</u><br>5.47 kcal/g | Nutritional composition N/R |
| Pritchard, 2014 (62) | UK<br>University students, staff and local community | BMI: M = 24<br>Gender: 10M, 23F | Adult | Within-subjects<br>Self-reported and researcher measured EI<br>EI measured for 1 day | N = 33 | No: Lunch only | <u>Lower ED</u><br>1 kcal/g<br><u>Higher ED</u><br>1.4 kcal/g | Nutritional composition not matched |
| Rolls, 2006 (63) | US<br>Local community | BMI: M = 22<br>Gender: 24F | Adult | Within-subjects<br>Researcher measured EI<br>EI measured for 2 days | N = 24 | Yes | <u>Lower ED</u><br>1.61 kcal/g<br><u>Higher ED</u><br>2.11 kcal/g | Nutritional composition not matched |
| Shide, 1995 (64) | US<br>Local community | BMI: M = 24<br>Gender: 48F | Adult | Within-subjects<br>Self-reported and researcher measured EI<br>EI measured for 1 day | N = 48 | No: Lunch food only | <u>Lower ED</u><br>0.46 kcal/g<br><u>Higher ED</u><br>1.02 kcal/g | Nutritional composition not matched |
| Silver, 2008 (65) | US<br>Adults aged over 60 in care | BMI: M = 24<br>Gender: | Adult | Within-subjects<br>Self-reported and researcher measured | N = 45 | No: Lunch only | <u>Lower ED</u><br>1.1 kcal/g<br><u>Higher ED</u> | Nutritional composition N/R |
|  | setting | 14M, 31F |  | EI<br>EI measured for 1 day |  |  | 2.2 kcal/g |  |
| Smethers, 2019 (66) | US<br>Children in childcare centre | BMI percentile:<br>M = 60<br>Gender:<br>26M, 23F | Children | Within-subjects<br>Self-reported and researcher measured EI<br>EI measured for 5 days | N = 49 | No: Breakfast, lunch entrée, dinner entrée and afternoon snack only | <u>Lower ED</u><br>1.47 kcal/g<br><u>Medium ED</u><br>1.82 kcal/<br><u>Higher ED</u><br>2.19 kcal/g | Nutritional composition N/R |
| Spill, 2011 (67) | US<br>Children in day care centre | BMI percentile:<br>M = 57<br>Gender:<br>18M, 21F | Children | Within-subjects<br>Researcher measured EI<br>EI measured for 1 day | N = 41 | No: Breakfast entrée, lunch entrees, dinner entrée, evening snack only only | <u>Lower ED</u><br>1.46 kcal/g<br><u>Medium ED</u><br>1.56 kcal/<br><u>Higher ED</u><br>1.95 kcal/g | Nutritional composition N/R |
| Stubbs, 1995a (18) | UK<br>Local community | BMI:<br>M = N/R<br>Gender:<br>6M | Adults | Within-subjects<br>Researcher measured EI<br>EI measured for 7 days | N = 6 | Yes | <u>Lower ED</u><br>1.15 kcal/g<br><u>Medium ED</u><br>1.34 kcal/<br><u>Higher ED</u><br>1.68 kcal/g | Nutritional composition not matched |
| Stubbs, 1995b (17) | UK<br>Local community | BMI:<br>M = N/R<br>Gender:<br>7M | Adults | Within-subjects<br>Researcher measured EI<br>EI measured for 14 days | N = 7 | Yes | <u>Lower ED</u><br>1.15 kcal/g<br><u>Medium ED</u><br>1.34 kcal/<br><u>Higher ED</u><br>1.68 kcal/g | Nutritional composition not matched |
| Stubbs, 1998a (15) | UK<br>Local | BMI:<br>M = 22 | Adults | Within-subjects<br>Researcher measured | N = 6 | Yes | <u>Lower ED</u><br>0.89 kcal/g | Nutritional composition matched |
|  | community | Gender:<br>6M |  | EI<br>EI measured for 14 days |  |  | <u>Medium ED</u><br>1.31 kcal/<br><u>Higher ED</u><br>1.76 kcal/g |  |
| Stubbs, 1998b (16) | UK (Scotland)<br>Local community | BMI:<br>M = 22<br>Gender:<br>6M | Adults | Within-subjects<br>Participant self-reported and researcher measured EI<br>EI measured for 14 days | N = 6 | Yes | <u>Lower ED</u><br>0.85 kcal/g<br><u>Higher ED</u><br>1.50 kcal/g | Nutritional composition matched |
| Tey, 2016 (68) | Singapore<br>University campus | BMI:<br>M = 22<br>Gender:<br>27M | Adults | Within-subjects<br>Participant self-reported and researcher measured EI<br>EI measured for 1 day | N = 27 | No: Lunch only | <u>Very low ED</u><br>0.27 kcal/g<br><u>Low ED</u><br>0.48 kcal/g<br><u>Medium ED</u><br>0.95 kcal/g<br><u>High ED</u><br>1.37 kcal/g<br><u>Very high ED</u><br>1.81 kcal/g | Nutritional composition not matched |
| Tey, 2018 (69) | Singapore<br>University campus | BMI:<br>M = 22<br>Gender:<br>32M | Adults | Within-subjects<br>Participant self-reported and researcher measured EI<br>EI measured for 1 day | N = 32 | No: Lunch food only | <u>Lower ED</u><br>0.11-0.13 kcal/g<br><u>Higher ED</u><br>0.49-0.5 kcal/g | Nutritional composition not matched |
| Westerstep, 1997(70) | Netherlands | BMI:<br>M = 22 & 27 | Adults | Within-subjects | N = 36 | No: Breakfast food and lunch only | <u>Lower ED</u><br>1.21 kcal/g | Nutritional composition not matched |
|  | University campus and local community | Gender: 36F |  | Participant self-reported and researcher measured EI<br>EI measured for 1 day |  |  | <u>Higher ED</u><br>1.78 kcal/g |  |
| Williams, 2013 (19) | US<br>University campus and local community | BMI: M = 26<br>Gender: 30M, 29F | Adults | Within-subjects<br>Researcher measured EI<br>EI measured for 1 day | N = 59 | No: Breakfast entrée, lunch entrée, dinner entrée only | <u>Lower ED</u><br>1.44 kcal/g<br><u>Higher ED</u><br>1.8 kcal/g | Nutritional composition matched for one comparison (water content manipulation), not matched for other comparisons (e.g. fat content manipulation) |

**Figure 1.**
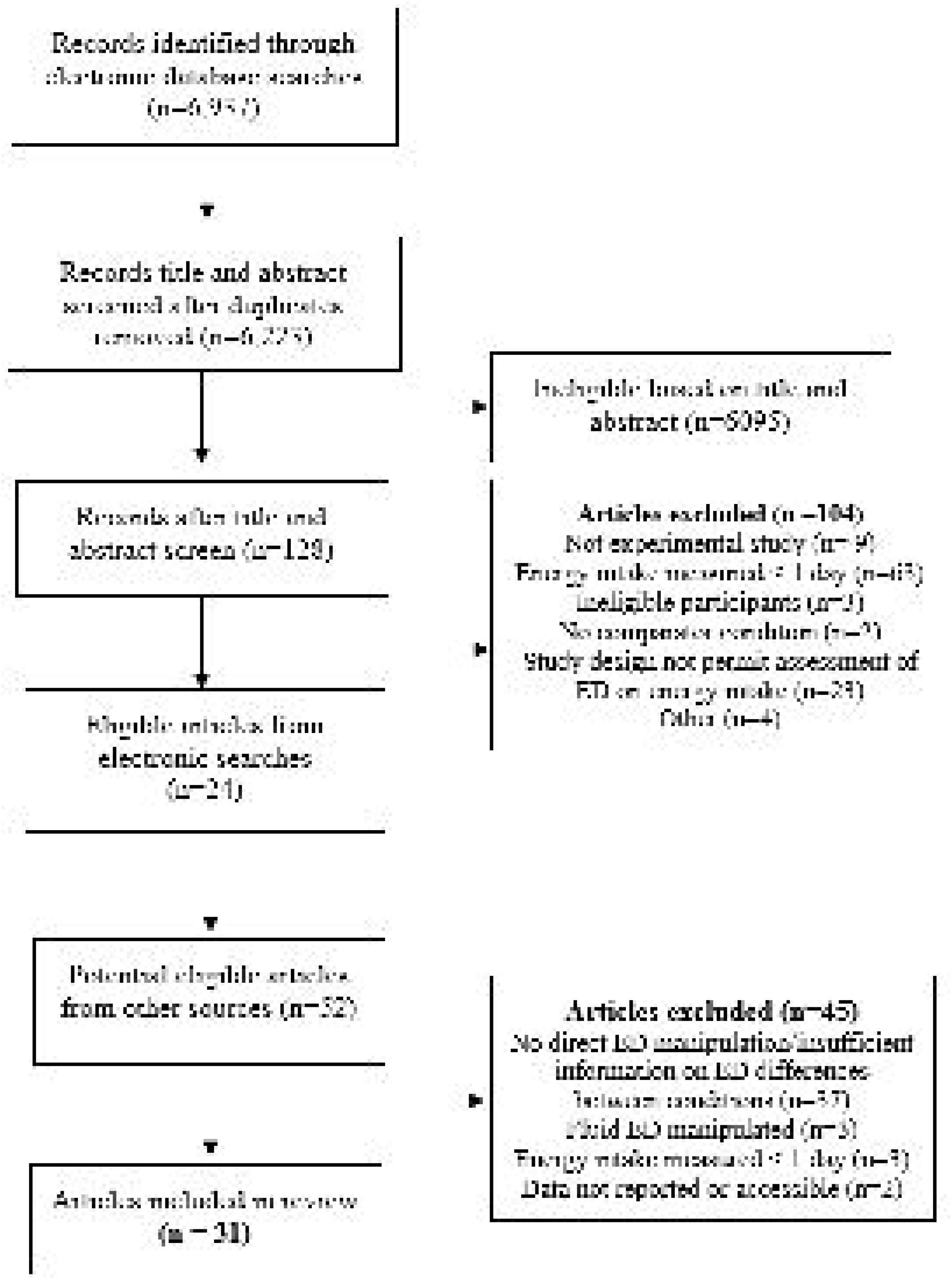
Systematic review study search and eligibility flowchart.

**Figure 2.**
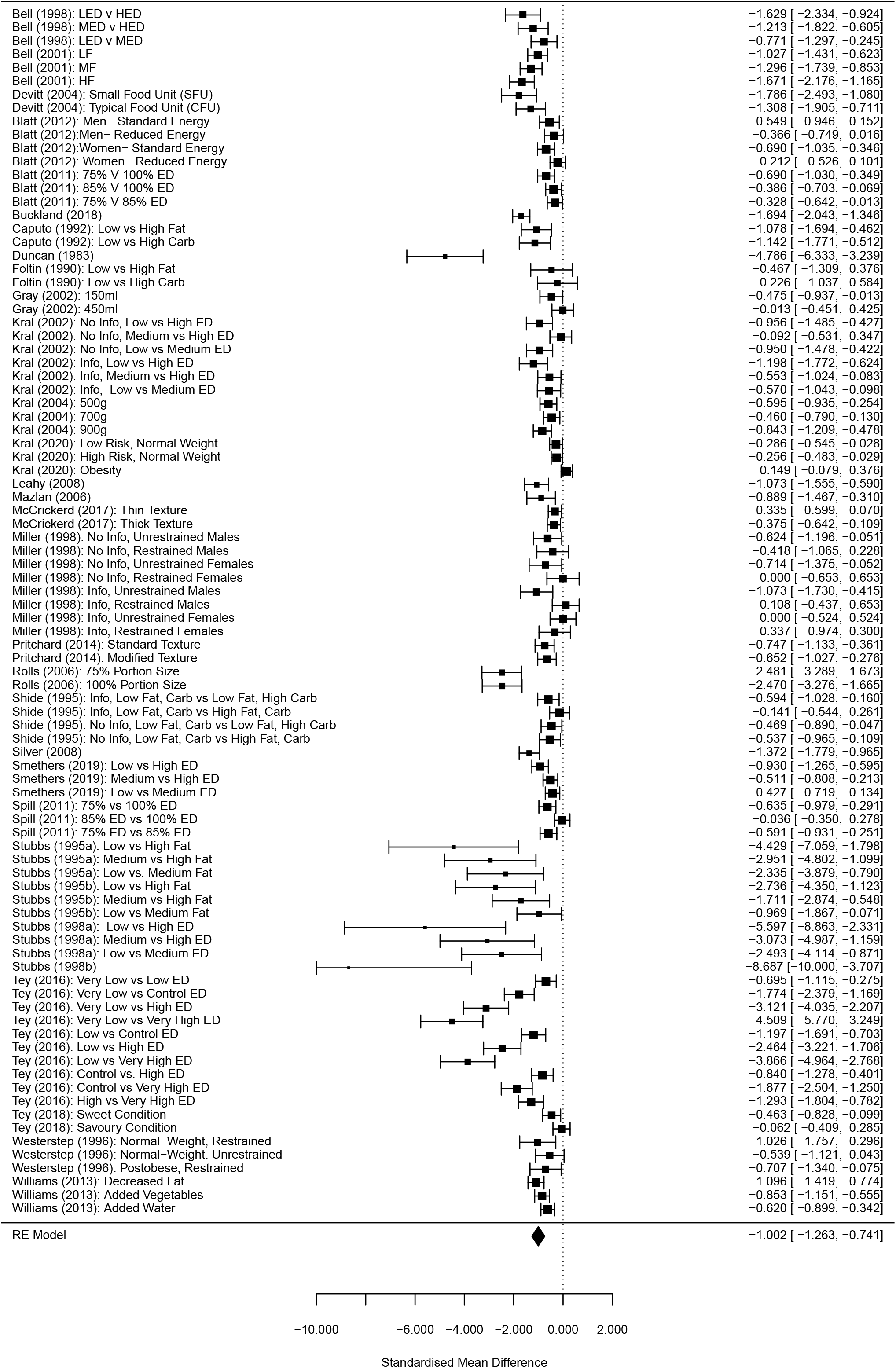
Forest plot of all studies included in primary meta-analysis.

### Risk of bias indicators

Of the thirty-one included studies, a sizeable minority (n=14) used participant self-report to quantify (in part or full) daily energy intake. Only n=2/31 studies did not report use of key participant exclusion criteria, n=2/31 studies did not report key methodological information, n=10/31 used either non-random allocation to energy density condition order or did not report on allocation method, n=15/31 required participants to eat at least one meal or test food in full as part of the procedure, in n=10/31 studies demand characteristics were not addressed (e.g. measurement of participant awareness of different energy density conditions). A minority of studies (n=5/31) had a small sample size (N<8), n=25/31 studies were not pre-registered and n=21/31 studies did not have a conflicts of interest statement or reported a relevant conflict. See supplementary online materials for individual study risk of bias information.

### Primary analyses

#### Effects on daily energy intake

The multi-level meta-analysis (90 effect sizes from 31 studies) indicated that there was a large effect of energy density condition on daily energy intake (SMD = -1.002 [95% CI: -0.745 to -1.266], Z = 7.54, p < .001, I^2^ = 92.1%), whereby serving lower energy dense foods was associated with lower daily energy intake. See Figure 1. Results remained significant in sensitivity analyses varying the size of within-subjects correlation for daily energy intake (see online supplementary material). Egger’s test was significant (Z = -10.82, p < .001), indicating possible publication bias, although Trim and Fill on a single level model identified 0 studies to be filled. See online supplementary materials for funnel plot. No DFBETAs were greater than >1, and leave-one-out analysis did not substantially influence the models (ps < .001). There were 28 effect sizes with confidence intervals which did not overlap the pooled analyses (outliers) and removing them from the analyses slightly reduced the pooled effect, but also the heterogeneity (SMD = -0.872 [95% CI: -1.001 to -0.742], Z = 13.21, p < .001, I^2^ = 60.6%).

#### Sub-group and meta-regression analyses on daily energy intake (outliers removed)

Sub-group analyses comparing adult vs. child and male vs. female samples were non-significant (see online supplementary materials). Moderation analysis comparing effects for which energy density was varied by manipulating nutritional composition (28 effect sizes) vs. kept constant (21 effect sizes) was non-significant (X^2^(1) = 1.00 p = .318), whereby energy density manipulations altering composition (SMD = -0.952 [95% CI: -0.694 to -1.209]) produced very similar effects on daily energy intake as those not altering composition (SMD = -0.859 [95% CI: -0.649 to -1.068]). Moderation analysis for number of meals/foods energy density was manipulated was statistically significant (X^2^(1) = 18.11, p < .001). Effects in which energy density of all foods served was manipulated (9 effects, SMD = -1.871 [95% CI: -1.313 to -2.430]) were associated with a larger impact on daily energy intake than effects in which not all food served was manipulated (53 effects, SMD = -0.796 [95% CI: -0.682 to - 0.910]). Meta-regression of the number of days energy intake indicated a negative but non-significant association with daily energy intake (b = -0.039 [95% CI: -.080 to 0.001], p = .060), whereby smaller effects on daily energy intake were associated with studies measuring energy intake for longer. All analysis results remained consistent with the inclusion of outliers from the primary model, although the meta-regression on length of study became significant (p = .018).

#### Linearity of relationship between manipulating energy density and daily energy intake (outliers removed)

The kcal/g of the highest energy density condition in each energy density comparison was not a significant predictor of effect on daily energy intake (b = -0.020 [95% CI: -0.129 to 0.089]) and effects for which both energy density conditions were < 1.75kcal/g vs. ≥1.75kcal/g in at least one condition produced similar sized results on daily energy intake (p = .160).

#### Risk of bias indicators (outliers removed)

Analyses examining whether effects of energy density on daily energy intake were dependent on whether studies used self-report vs. measured energy intake, addressed demand characteristics and conflicts of interest vs. did not were all non-significant. Studies which either did not use or failed to report on random allocation to energy density conditions tended to produce larger effects on daily energy intake than studies which did report use of random allocation (p = .045), but both types of study were individually significant. See online supplementary materials for results in full.

#### Analyses limited to studies manipulating energy density of all foods/meals

There was a large effect of energy density condition on daily energy intake (kcal difference between higher and lower energy density conditions = -855.85 [95% CI: -616.18 to -1095.52], Z = 7.00, p < .001, I^2^ = 97.4%). See Figure 3. Removal of outliers (5 effects) slightly reduced the kcal difference (−709.01 [95% CI: -602.04 to –815.97], Z = 12.99, p < .001, I^2^ = 85.4%). There was a significant association between difference in energy density between meals and differences in kcals consumed between conditions/meals (b = -1510.70 [95% CI: -1236.08 to -1785.33], p < .001) and with outliers removed the association was smaller but remained significant (b = - 309.31 [95% CI: -115.91 to -502.71], Z = 3.13, p = .002). Length of study (number of days) was not significantly related to effects on daily energy intake and analyses examining non-linearity were also non-significant, as in the primary analyses (see online supplementary materials).

**Figure 3.**
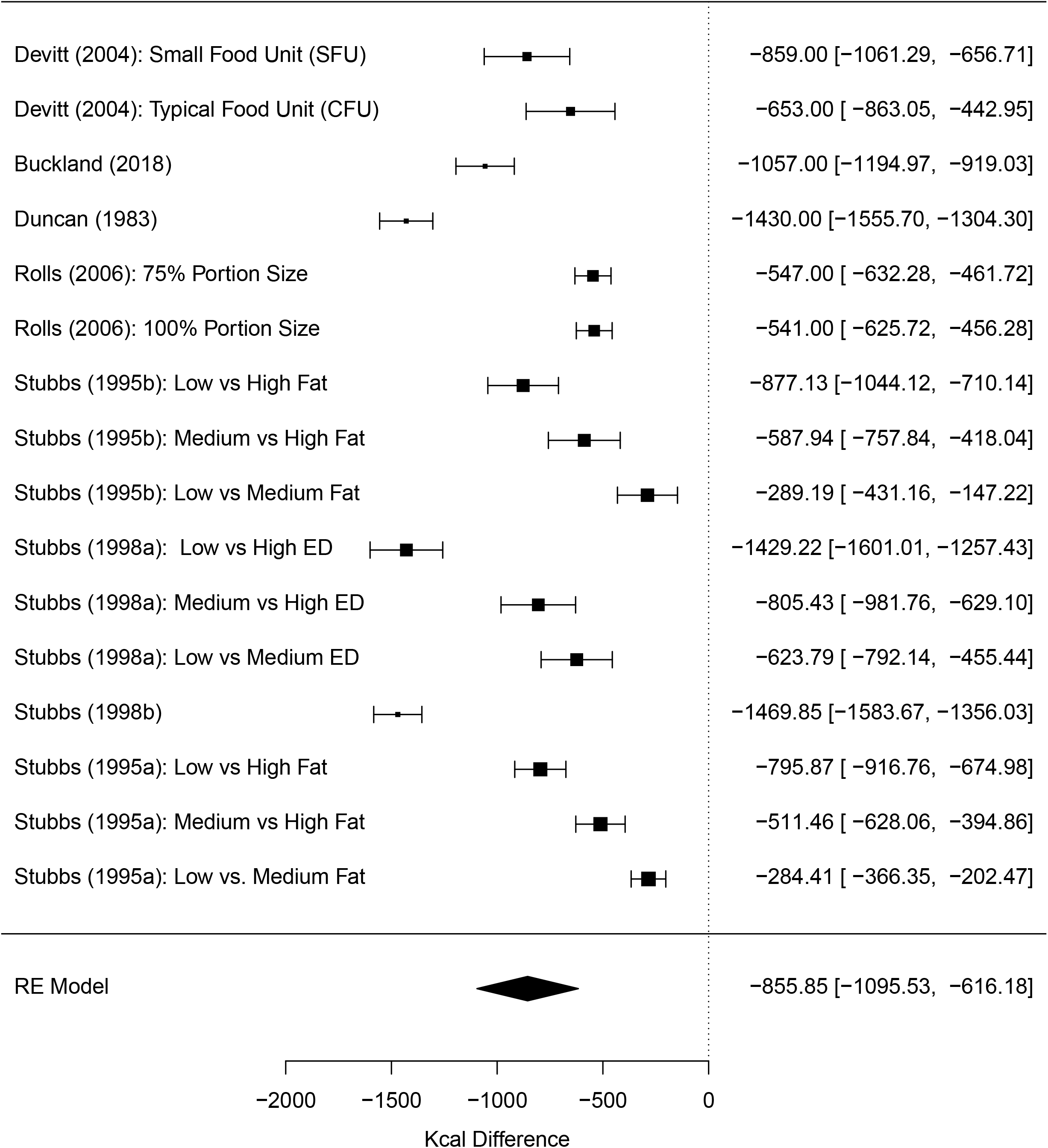
Forest plot for analyses limited to studies which manipulated energy density for all foods served.

#### Analyses limited to studies not manipulating energy density of all foods/meals

There was a large effect of energy density condition on daily energy intake (kcal diff = -237.84 [95% CI: - 148.13 to -327.54], Z = 5.20, p < .001, I2 = 95.9). See Figure 4. Twelve outliers were identified, and removal of these effects slightly reduced the kcal difference, but also the heterogeneity (−208.17 [95% CI: -160.00 to -256.37], Z = 8.47, p < .001, I2 = 75.5%). There was a significant association between difference in energy density between conditions and the difference in daily energy intake between conditions (b = -331.86 [95% CI: -234.62 to - 429.13], Z = 6.69, p < .001) and with outliers excluded, the effect remained significant but somewhat smaller (b = -104.50 [95% CI: -12.03 to 196.98], Z = 2.21, p = .027). Length of study was not significantly related to effects on daily energy intake. For analyses examining potential non-linearity, results were largely consistent with the primary analyses. See online supplementary materials.

**Figure 4.**
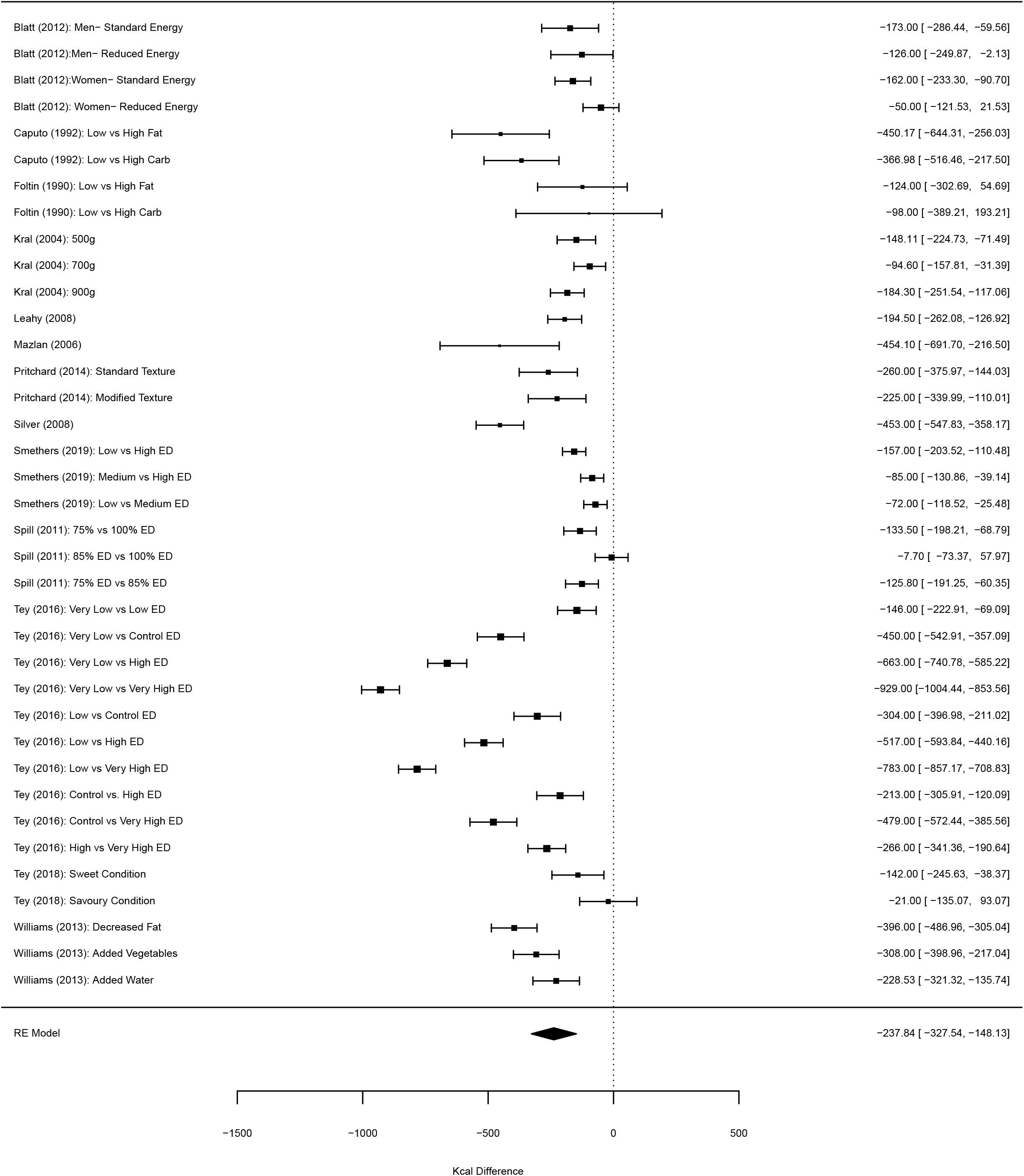
Forest plot for analyses limited to studies which manipulated energy density of some but not all foods served.

#### Difference in total kcals served and daily energy intake

Among studies that did not manipulate energy density of all foods/meals, there was a significant association between differences in kcals served between energy density conditions and difference in daily energy intake (b = -0.774 [95% CI: -0.644 to -0.905], Z = 11.64, p < .001), whereby a 100kcal difference in energy served (due to energy density manipulation) was predictive of a 77kcal difference in daily energy intake. Results remained significant with removal of outliers. See online supplementary materials.

#### Energy intake during manipulated energy density meals vs. later in the day

For studies that provided complete data (i.e. Ms and SDs) on both energy intake from energy density manipulated foods/meals and energy intake from subsequent non-manipulated foods/meals (16 effects from 7 studies), the difference in kcals consumed from manipulated meals between higher and lower energy density conditions was - 330.78kcals ([95% CI: -224.27 to - 437.29), Z = 6.09, p < .001, I^2^ = 100%) and similar in sensitivity analyses that varied size of SD for manipulated meals that required compulsory eating [-326.40 ([95% CI:-222.53 to - 431.31]. There was a small increase in kcals consumed after consuming lower vs. high energy dense food, but this increase was not statistically significant (kcals = 35.08 [95% CI: - 28.32 to 98.48], Z = 0.28, p = .278, I^2^ = 95.15).

#### Body weight

Pooled across the five studies that provided data on weight change, weight loss tended to be greater in lower compared to higher energy dense conditions, but this difference was not statistically significant, kg change = -0.69 [95% CI: -1.43 to 0.04). See Figure 5. Results were similar in sensitivity analyses. See online supplementary materials.

**Figure 5.**
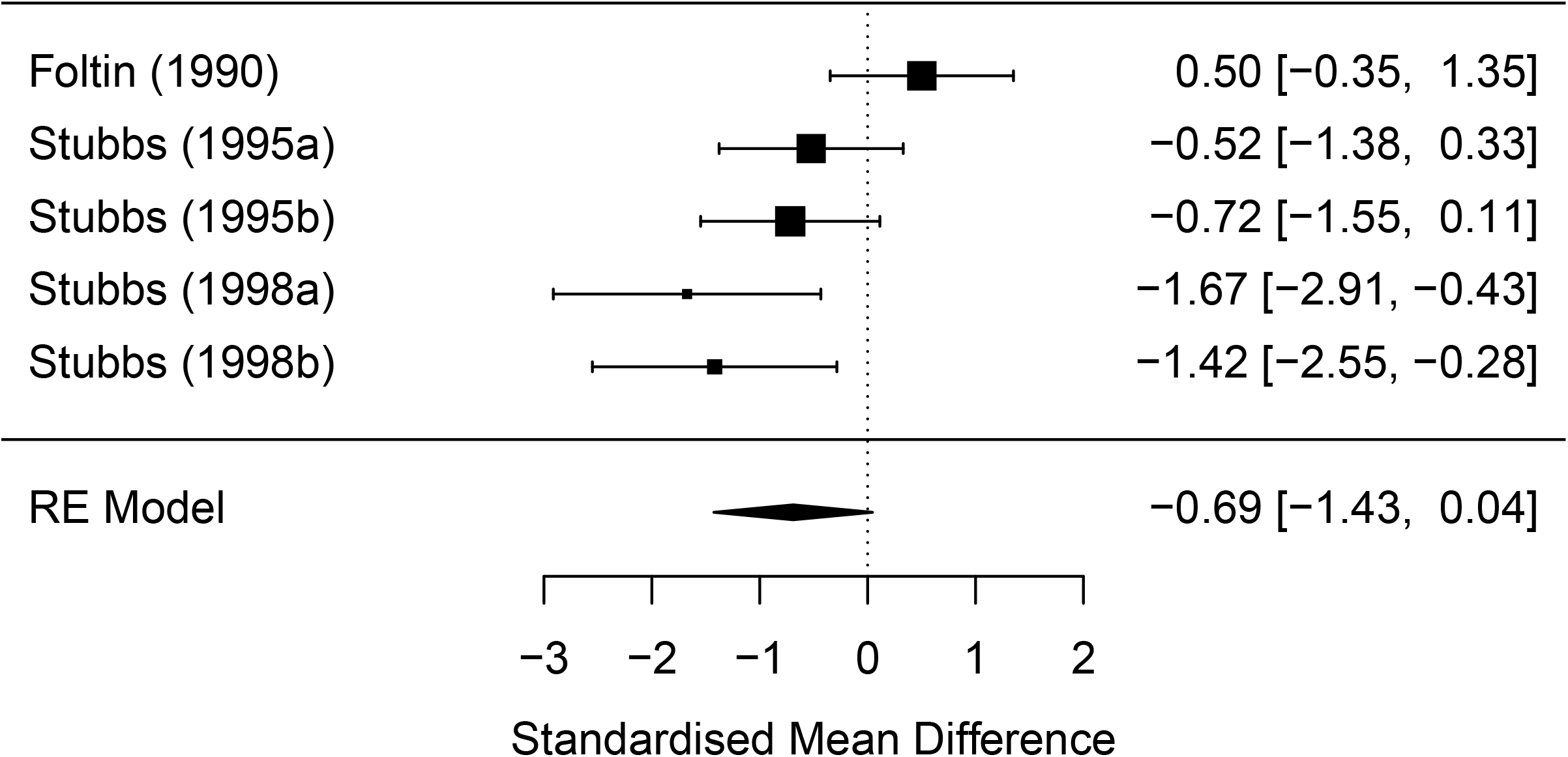
Body weight meta-analysis forest plot.

## Discussion

Serving lower energy dense (vs. higher energy density) foods significantly reduced daily energy intake and this effect was statistically large. Studies with the most pronounced differences in energy density produced the largest changes to daily energy intake (i.e. a dose-dependent response) and studies which manipulated the energy density of all foods served produced larger effects on daily energy intake than studies that did not manipulate all foods. The impact energy density had on daily energy intake was similar among males vs. females and in adults vs. children. Studies tended to manipulate energy density by either altering macronutrient content (e.g. by reducing %kcals from fat) or by holding macronutrient content (e.g. by increasing water content) and both manipulations produced similar sized effects on daily energy intake. Due to the available data, we were unable to examine whether distinct macronutrient manipulations (e.g. replacing fat with protein as opposed to carbohydrate) affected daily energy intake differentially. However, over and above absolute changes to energy density, any nutrient specific effects may be relatively subtle (14). Due to the satiety-providing effects of protein, decreasing energy by increasing protein content may have a more pronounced effect on daily energy intake (38, 39), but evidence is mixed (40, 41).

We found consistent evidence that the relationship between energy density and daily energy intake was strong and linear. Consistent with this, among studies that manipulated energy density for some but not all food served, analyses suggested that for every 100 fewer kcals of food served to participants (due to reduced energy density), daily energy intake was reduced by approximately 77kcals. Furthermore, in these studies participants consumed approximately 326kcals fewer during the lower (vs. higher) energy dense meals but increased their later ad-libitum energy intake (from non-manipulated foods) by a non-significant 35kcals. Therefore, unlike manipulation of food product portion size for which more substantial compensation appears to occur (37), there is minimal evidence of energy intake compensation in response to manipulations of food energy density. An implication of the present findings is that public health policies which reduce energy density of food being sold (e.g. through voluntary industry reformulation or mandatory action) are likely to be more effective in reducing daily energy intake than policies which target portion size alone.

A limitation of included studies was their relatively short duration (between 1 and 14 days). We found some inconsistent evidence that the length of time energy intake was measured for moderated findings, whereby effects of energy density on daily energy intake were smaller among studies with longer duration in our main analysis. However, the statistical significance of this effect was dependent on the exclusion of outliers from analyses and this association was not observed when studies that manipulated all foods (vs. did not) were analysed separately. This finding may indicate that over time consumers learn about the energy density of food served and adapt their food intake, but this adaptation is only partial. A small sub-set of studies examined change in body weight and although after being served lower vs. energy dense foods participants tended to lose more weight (1.4kg difference), this difference was not statistically significant. Although previous studies that have directed participants to reduce energy density through dietary advice provide evidence for significant changes to body weight (1, 25), the effect of reformulating the energy density of foods on body weight therefore remains less clear. Future research will therefore be needed examining the effect that manipulations of energy density have on body weight in order to understood whether mass reformulation of the energy density of food products is likely to benefit population level obesity.

Contrary to suggestions that humans may be more sensitive to changes in energy density to less energy dense foods (23, 24), we found no evidence that the impact of reducing energy density of food served was non-linear in nature; studies comparing two low energy density conditions (e.g. 1.1kcal/g vs. 0.8kcal/g) produced similar sized effects to studies comparing more energy dense foods (e.g. 2.6kcal/g vs. 2.3kcal/g). However, the majority of studies examined lower food energy densities, as opposed to ‘highly’ energy dense foods (i.e. ≥4kcal/g) and it may be the case that differences would be observed for the latter. Further research directly addressing this question will now be important because public health approaches would presumably target reformulation of highly energy dense foods, as opposed to food products that are already relatively low in energy density.

There are strengths and limitations to the present research. We followed best practice guidelines for systematic review methodology and attempted to identify eligible published and unpublished articles using a combination of supplementary methods including grey literature searching and contact authors of eligible articles. Methodological quality of included studies was variable, but studies tended to be well-reported, few were of very small sample size and most study designs addressed demand characteristics. We assessed whether a range of potential risk of bias indicators affected results in sub-group analyses and found little convincing evidence that risk of bias indicators predicted study outcomes. As discussed, study durations were relatively short and the artificial nature of the laboratory settings used in most studies increases confidence in experimental control but at the expense of ecological validity (32, 42). It may be the case that alterations to food energy density would be associated with greater compensation outside of the laboratory when concerns about social desirability are reduced and/or a wider selection of food is available (32, 43, 44), which would result in smaller effects on daily energy intake and body weight. It is also important to note that a number of included studies allowed participants to consume foods and meals outside of the laboratory and later self-report this intake, and in these studies the effect of manipulating energy density of food served in the laboratory on daily energy intake was still sizeable. As noted, the relatively short duration of studies is a limitation and it may be that over longer time periods, the post-ingestive consequences of lower energy density foods would result in dietary learning. However, it is not clear how long foods would need to be consumed for, as in one study repeated daily exposures of higher vs. lower energy density versions of the same product for 5 days produced no evidence of dietary learning (45). It should also be noted that we detected evidence of funnel plot asymmetry which may be indicative of publication bias. However, this appears to have been largely caused by there being a number of studies that had particularly large manipulations to energy density which would be expected to cause large decreases in daily energy intake and therefore contribute to asymmetry. A final limitation was that we were only able to examine a small number of participant characteristics in moderation analyses (sex, age) and it may be the case that there are other characteristics (e.g. BMI, socioeconomic status) or participant traits (e.g. satiety responsiveness) that moderate the effect reducing energy density has on daily energy intake.

### Conclusions

Experimental studies indicate that decreasing energy density of food products has a strong and largely linear effect on daily energy intake, although effects on body weight are less clear and warrant further study. Reformulation of the energy density of food products may be an effective public health approach to reducing population level energy intake.

### Studies included in review

Refs 15-20, 46-70.

## Data Availability

Data described in the manuscript, code book, and analytic code is publicly and freely available without restriction at https://osf.io/dj4yf/

## Acknowledgements

N/A.

## Funding

No external funding.

## Conflicts of Interest

All authors report no conflicts of interest. ER has previously received funding from the American Beverage Association and Unilever for projects unrelated to the present research.

## Author Contributions

ER designed the research, conducted the research, had primary responsible for the final content and wrote the paper. AJ designed the research, conducted the research, analysed data and wrote the paper. IML, MK and ZP conducted the research.

## Notes

### Funding Statement

This study did not receive any funding

