## Supplementary material for "Calorie reformulation: A systematic review and meta-analysis examining the effect of manipulating food energy density on daily energy intake and body weight": Online supplementary material

Online Supplementary Materials

Search terms used

PubMed, SCOPUS, Psych Info: (Energy density OR portion size OR food reformulation) AND (energy intake OR calories OR food intake OR appetite OR eating). Filter: Humans (applied in PubMed and Psych Info).

Deviations from planned analyses

We had originally planned to systematically review and meta-analyse studies that had manipulated portion size of foods and studies that had manipulated energy density of foods in the same project. However, after conducting searches and identifying eligible articles we determined that the scope of the review is too large to be completed in line with the original project timeline, as a result of staffing during the COVID pandemic and a larger number of eligible energy density studies than anticipated. For these reasons, we separated out portion size and energy density studies into two distinct projects. This decision was made during piloting of data extraction. The only major change to the pre-registered protocol was the exclusion of studies that manipulated portion size of food (and not energy density) and therefore a planned analysis examining the effect of ‘study type’ (i.e. portion size vs. energy density) was not conducted. We also made minor deviations from the planned protocol (e.g. separate analyses of studies that manipulated energy density of all foods during the day vs. no) and they are described in the main manuscript.

Planned influential case and publication bias analyses

We characterised outliers as any effect sizes for which the upper bound of the 95% confidence interval was lower than the lower bound of the pooled effect confidence interval (i.e., extremely small effects) or for which the lower bound of the 95% confidence interval was higher than the upper bound of the pooled effect confidence interval (i.e., extremely large effects). If any outliers were identified, then we planned to report the results of meta-analyses with the outliers removed. To address influential cases, we computed DFBETAS values for each effect size. Influential cases were identified if DFBETAS values > 1 (indicative of a >1 change in the standard deviation of the estimated co-efficient after removal of the study) (1). To increase sensitivity, we conducted leave-one-out analyses by removing each study (k) from the analyses and refitting the model. If the removal of k substantially influences the model (statistical significance of the model changes from p < .05 to p > .05 (or p >.05 to p < .05), this was classed as an influential case. We examined evidence for publication bias in the primary analyses by examining asymmetry of effect sizes. We plotted and visually inspected funnel plots. Next, we conducted an Egger’s test of asymmetry and Trim and Fill procedure. For Egger’s test (2), if the intercept is significantly different from 0 at p >.10 this is indicative of bias. Trim and Fill (3) removes less precise studies which might case any asymmetry (‘trim’), re-estimates the overall effect size, and then replaces removed studies and missing counterparts (‘fill’) required for symmetry. We planned to report i) the number of missing (‘filled’) studies, and ii) the estimate of the effect size following their inclusion.

Additional Analyses

*Primary multi-level model imputation of different within-subjects correlation coefficients.* Imputation of correlations of r = 0.6 and r = 0.4 (as opposed to 0.8) for the within-subjects correlation for daily energy intake reduced the statistical magnitude of the effect but not the statistical significance [SMD = -0.744 [95% CI: -0.547 to -0.942] and SMD = -0.621 [95% CI: -0.454 to -0.787], respectively.

*Sub-group analyses from primary model (outliers excluded)*

*Age group*: Moderation analysis for adults (57 effect sizes) vs children (5 effect sizes) was not statistically significant (X2(1) = 0.40, p = .526). For adults the SMD = -0.892 [95% CI: -1.039 to -0.746], and for children the SMD = -0.713 [95% CI: -0.521 to -0.905].

*Sex*: Moderation analyses for males (23 effect sizes) vs females (23 effect sizes) vs mixed samples (16 effect sizes) was not statistically significant (X2(2) = 0.27, p = .875). For males the SMD = -0.979 [95% CI: -0.595 to -1.362], for females the SMD = -0.792 [95% CI: -0.613 to -0.971], and for mixed the SMD = -0.907 [95% CI: -0.713 to -1.001].

*Risk of bias indicators from primary model (outliers excluded)*

*Random allocation used:* Moderation analysis of random allocation (20 effect sizes) vs non-random allocation (42 effect sizes) was statistically significant (X2(1) = 4.03, p = .045). For effects using random allocation the SMD = -0.676 [95% CI: -0.549 to -0.805], for non-random allocation the SMD = -0.983 [95% CI: -0.805 to -1.162].

*Energy intake measure:* Moderation analysis of objective researcher measured (37 effect sizes) vs self-report based measures of energy intake (25 effect sizes) was not significant (X2(1) = 1.40, p = .237). For objective researcher measurement the SMD = -0.956 [95% CI: -0.763 to -1.150], for self-report measurement the SMD = -0.792 [95% CI: -0.603 to -0.981].

*Demand characteristics:* Moderation analysis of effects in which demand characteristics were addressed (42 effect sizes) vs not addressed (20 effect sizes) was not significant (X2(1) < 0.01, p < .931). For demand characteristics addressed the SMD = -0.869 [95% CI: -0.705 to -1.033], if demand characteristics were not addressed the effect size was SMD = -0.879 [95% CI: -0.656 to -1.102].

*Conflicts of interest:* Moderation analyses of effects in which conflicts of interest (21 effect sizes) were reported on and absent vs not reported or relevant interests declared (41 effect sizes) was not significant (X2(1) = 1.60, p = .205). If conflict of interests were reported on and absent the SMD = -0.779 [95% CI: -0.619 to -0.939], if conflicts of interests were not reported (or a relevant interest declared) the SMD = -0.959 [95% CI: -0.758 to -1.160].

*Analyses limited to studies manipulating energy density of all foods/meals*

*Kcal/g for higher ED meals (meta-regression):* No significant association with effect of energy density on daily energy intake (p = .984). Without outliers (p = .257) results remined the same.

*1.75 kcal/g sub-group analysis*: Moderation was not significant (p = .993). Without outliers (p = .454) results remained the same.

*Number of days energy intake measured (meta-regression):*	No significant association with effect of energy density on daily energy intake (p = .779) and resulted remained the same without outliers (p = .733).

*Analyses limited to studies not manipulating energy density of all foods/meals*

*Number of days energy intake measured (meta-regression)*: No significant association with outliers included (p = .776), or excluded (p = .568).

*1.75 Kcal/g sub-group analysis*: Moderation was not significant with outliers included (p = .557), or excluded (p = .267).

*Kcal/g for higher ED meals (meta-regression):* There was a significant association between Kcal/g for the higher meal and the reduction in kcals between conditions (b = -128.02 [95% CI: -22.53 to -233.50], Z = 2.38, p = .017). However, when outliers were removed, the association was substantially smaller and no longer significant (b – 54.03 [95% CI: -109.42 to 1.37], Z = 1.91, p = .056).

*Difference in total kcals served and daily energy intake*. Among studies that did not manipulate energy density of all foods/meals, there was a significant association between differences in kcals served between energy density conditions and difference in daily energy intake (b = -0.774 [95% CI: -0.644 to -0.905], Z = 11.64, p < .001). If outliers were excluded the association was still significant (b = -.257 [95% CI: -0.095 to -0.418], Z = 3.11, p = .002).

*Effect of compulsory vs. no compulsory eating.* No significant moderation effect of compulsory vs no compulsory eating (X^2^(1) = 0.182, p = .670). If outliers were excluded the association remained non-significant (X^2^(1) = 0.004, p = .948)

Funnel Plots

*Figure S1. Funnel plot of all effects from primary model. X axis is standardised mean difference.*

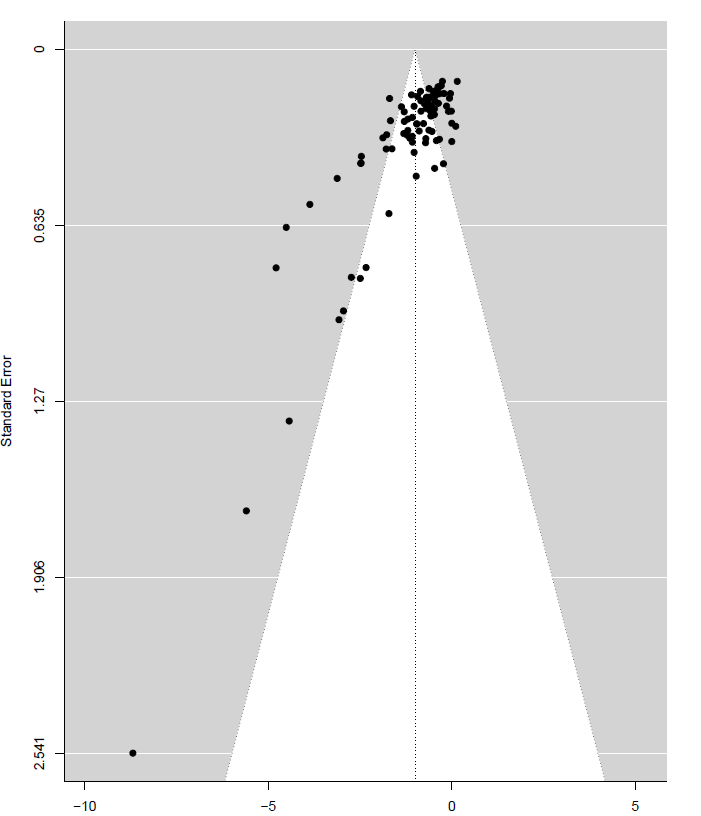

*Figure S2. Funnel plot of effects included in model examining studies that manipulated energy density of all foods/meals. X axis is difference in daily kcals consumed.*

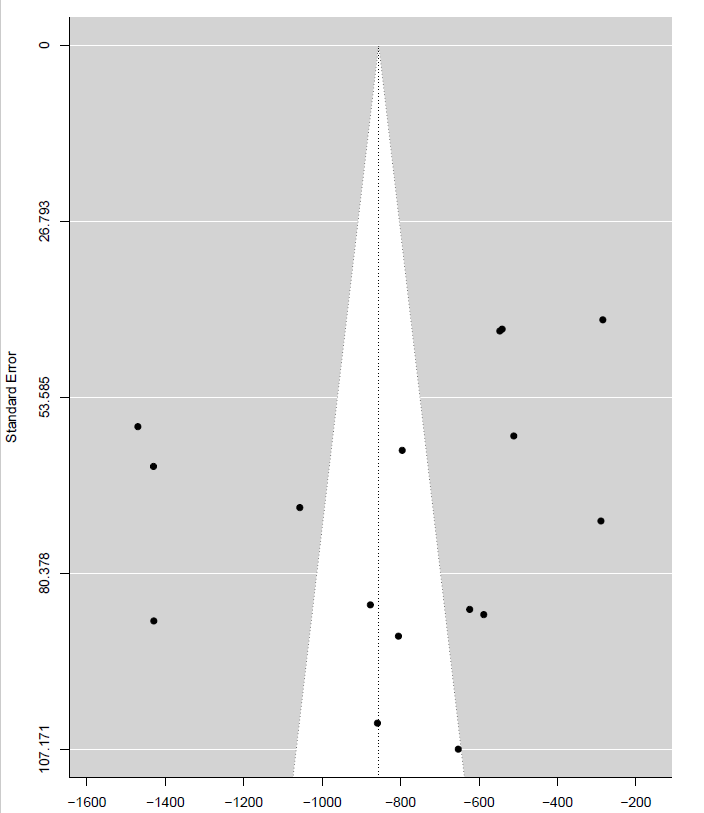

*Figure S3. Funnel plot of effects included in model examining studies that manipulated energy density of some but not all foods/meals.* *X axis is difference in daily kcals consumed.*

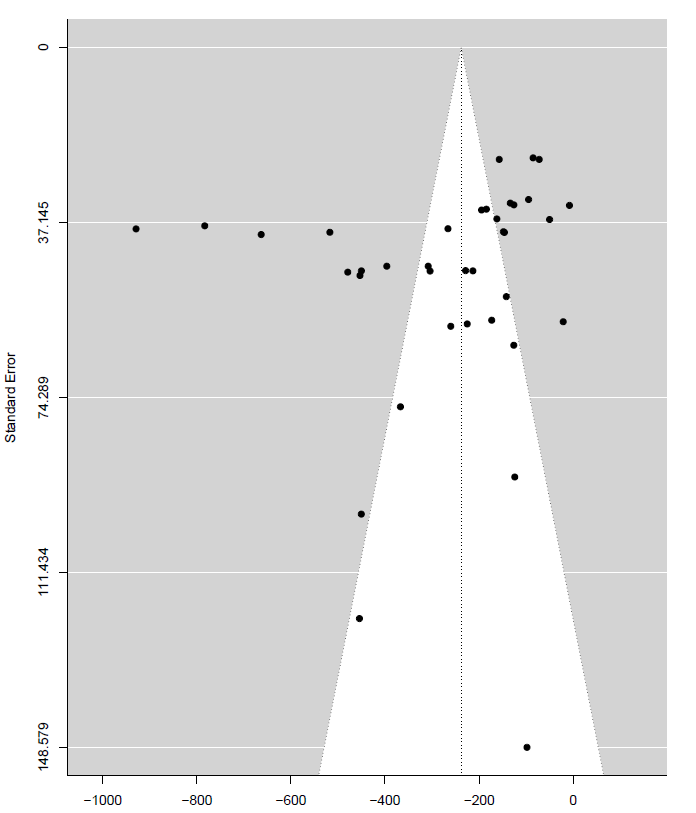

Body weight sensitivity meta-analyses

In sensitivity analyses we varied the imputed SDs of mean weight change in the lower and higher energy density condition from the 150% value used in our main analyses to 100% (Figure S4) and 200% (Figure S5). Results were consistent with the main analyses, a small non-significant difference was observed, whereby lower energy density conditions were associated with greater negative weight change compared to the higher energy density conditions.

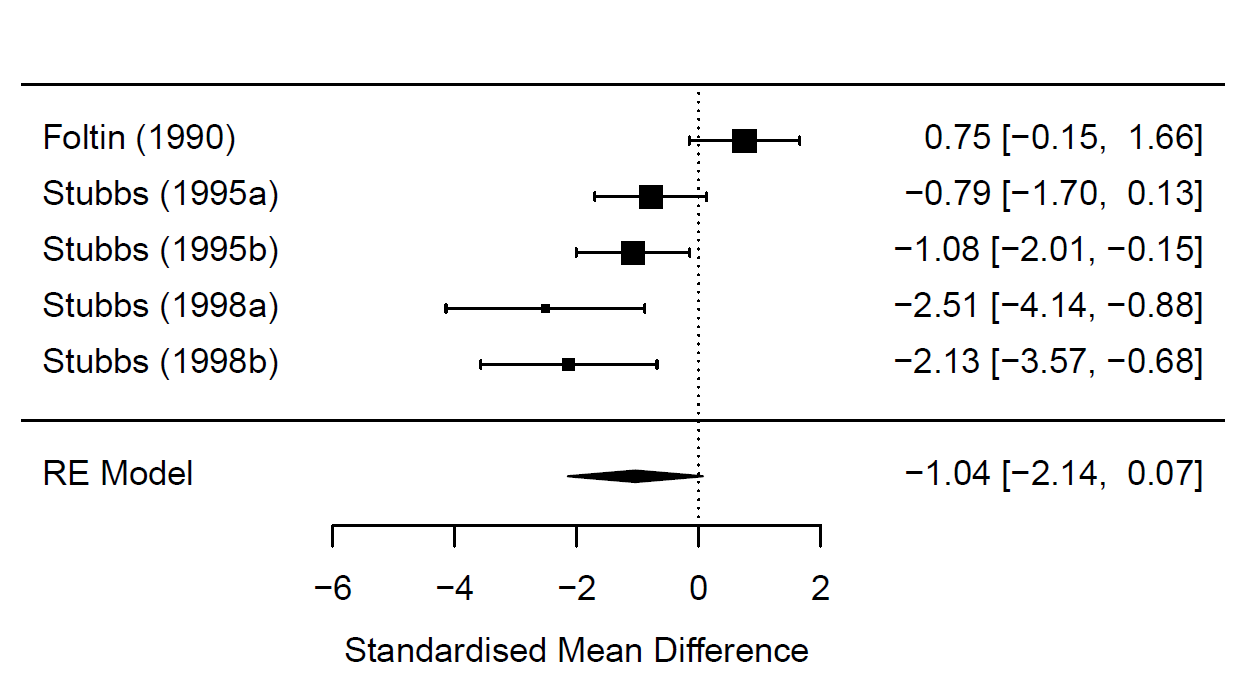
*Figure S4. Body weight meta-analysis, SDs imputed as 100% of mean weight change*

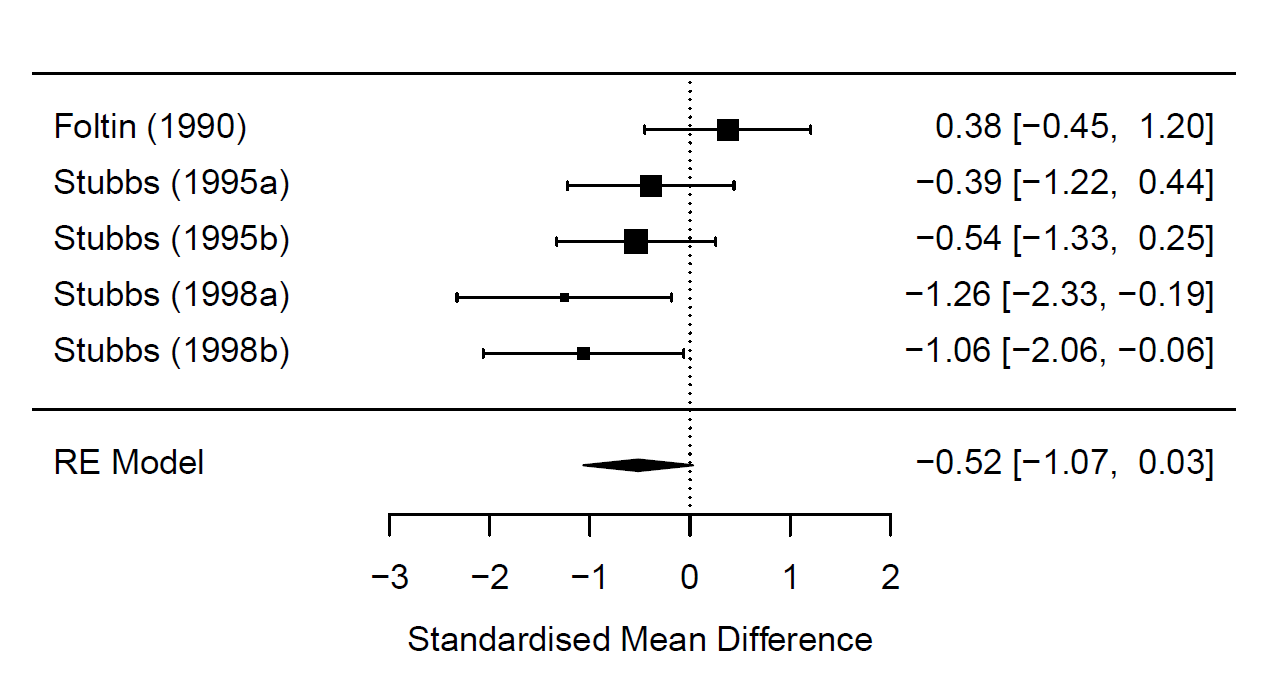
*Figure S5. Body weight meta-analysis, SDs imputed as 200% of mean weight change*

Supplementary table 1. Risk of bias information for included studies

| **Study** | **Self-reported energy intake used** | **Key exclusion criteria omitted or unclear** | **Lack of key method information** | **No/unclear random allocation to conditions** | **Required to consume meal(s) in full** | **Demand characteristics not addressed** | **Small sample size** | **Study was not pre-registered** | **CIs missing or reported** |
| --- | --- | --- | --- | --- | --- | --- | --- | --- | --- |
| Bell, 1998 | N | N | N | N | Y | N | N | Y | Y |
| Bell, 2001 | N | N | N | N | Y | N | N | Y | Y |
| Blatt, 2011 | N | N | N | N | N | N | N | N | N |
| Blatt, 2012 | N | N | N | Y | Y | N | N | N | Y |
| Buckland, 2018 | Y | N | N | N | Y | Y | N | Y | Y |
| Caputo, 1992 | Y | Y | Y | Y | Y | N | N | Y | Y |
| Devitt, 2004 | N | N | N | N | N | N | N | Y | Y |
| Duncan, 1983 | N | N | N | Y | N | N | N | Y | Y |
| Foltin, 1990 | Y | N | N | Y | Y | N | Y | Y | Y |
| Gray, 2002 | Y | N | N | Y | Y | N | N | Y | Y |
| Kral, 2002 | N | N | N | Y | Y | Mixed | N | Y | Y |
| Kral, 2004 | N | N | N | Y | Y | N | N | Y | N |
| Kral, 2020 | Y | N | N | N | Y | Y | N | Y | Y |
| Leahy, 2008 | N | N | Y | Y | N | Y | N | Y | N |
| Mazlan, 2006 | N | N | N | N | Y | N | N | Y | Y |
| McCrickerd, 2017 | Y | N | N | N | N | N | N | Y | Y |
| Miller, 1998 | Y | N | N | N | N | Mixed | N | Y | Y |
| Pritchard, 2014 | Y | N | N | N | N | N | N | Y | N |
| Rolls, 2006 | N | N | N | Y | N | N | N | Y | N |
| Shide, 1995 | N | N | N | Y | Y | N | N | Y | Y |
| Silver, 2008 | Y | N | N | N | N | Y | N | Y | Y |
| Smethers, 2019 | Y | N | N | N | N | Y | N | N | N |
| Spill, 2011 | N | N | N | N | N | Y | N | N | N |
| Stubbs 1995a | N | N | N | N | N | N | Y | Y | Y |
| Stubbs 1995b | N | N | N | N | N | Y | Y | Y | Y |
| Stubbs, 1998a | N | N | N | N | N | Y | Y | Y | Y |
| Stubbs, 1998b | Y | Y | N | N | N | N | Y | Y | Y |
| Tey, 2016 | Y | N | N | N | Y | N | N | N | N |
| Tey, 2018 | Y | N | N | N | Y | Y | N | N | N |
| Westerstep, 1997 | Y | N | N | N | Y | Y | N | Y | Y |
| Williams, 2013 | N | N | N | N | N | N | N | Y | N |
